# Evaluation of Dreem headband for sleep staging and EEG spectral analysis in people living with Alzheimer’s and older adults

**DOI:** 10.1101/2024.12.18.24319240

**Authors:** Kiran K G Ravindran, Ciro della Monica, Giuseppe Atzori, Hana Hassanin, Ramin Nilforooshan, Victoria Revell, Derk-Jan Dijk

## Abstract

**Introduction:** Portable electroencephalography (EEG) devices offer the potential for accurate quantification of sleep at home but have not been evaluated in relevant populations.

**Methods:** We assessed the Dreem headband (DHB), and its automated sleep staging algorithm in 62 older adults [Age (mean±SD) 70.5±6.7 years; 12 Alzheimer’s]. The accuracy of sleep measures, epoch-by-epoch staging, and the quality of EEG signals for quantitative EEG (qEEG) analysis was compared to standard polysomnography (PSG) in a sleep laboratory.

**Results:** The DHB algorithm accurately estimated total sleep time (TST) and sleep efficiency (SEFF) with a Symmetric Mean Absolute Percentage Error (SMAPE) <10%. Wake after sleep onset (WASO) and number of awakenings (NAW) were underestimated (WASO: ∼17 minutes; NAW: ∼9 counts) with SMAPE <20%. Sleep onset latency (SOL) was overestimated by ∼30 minutes when using the entire DHB recording period, but it was accurate (Bias: 0.3 minutes) when estimated over the lights-off period. Stage N3 and total non-rapid eye movement (REM) sleep durations were estimated accurately (Bias <20 minutes), while REM sleep was overestimated (∼25 minutes; SMAPE: ∼24%). Epoch-by-epoch sleep/wake classification showed acceptable performance (MCC=0.77±0.17) and 5-stage sleep classification was moderate (MCC=0.54±0.14). After artefact removal, 73% of the recordings were usable for qEEG analysis. Concordance (p<0.001) of EEG band power ranged from moderate to good: slow wave activity r^2^=0.57; theta r^2^=0.56; alpha r^2^=0.65; sigma power r^2^=0.34.

**Conclusion:** DHB algorithm provides accurate estimates of several sleep measures and qEEG metrics. However, further improvement in REM detection is needed to enhance its utility for research and clinical applications.

**Statement of Significance:** Wearable electroencephalography (EEG) devices such as the Dreem headband (DHB), hold promise for accurate monitoring of sleep at home and improving our understanding of neurodegenerative conditions. This study is important because it provides a comprehensive evaluation of the DHB in older adults, including people living with Alzheimer’s. The DHB automatic sleep staging algorithm demonstrated good concordance with polysomnography for most standard sleep statistics, and epoch-by-epoch sleep/wake classification. A novel aspect of the study is the evaluation of the suitability of the DHB EEG signal for quantitative EEG analysis. Our findings highlight the DHB’s key strengths and provide critical recommendations for improving usability and performance to establish its potential utility for large-scale, objective sleep monitoring in community-dwelling older adults.

## Introduction

Disturbances of sleep and circadian rhythms are reported to be highly prevalent in neurodegenerative conditions and negatively impacts the quality of life of people living with dementia (PLWD) and their caregivers [1], [2], [3], [4]. Accurate monitoring of sleep structure and timing can contribute to detecting early signs of neurodegeneration, monitoring disease progression in PLWD, and assessing of the effectiveness of interventions to improve sleep quality [5]. Clinical polysomnography (PSG), the gold standard for sleep measurement, allows for high-quality sleep characterization but comes at considerable cost, is burdensome to the individual, requires expert oversight, and is not suitable for longitudinal sleep monitoring.

In response to these limitations of PSG, consumer sleep technologies are being increasingly adopted for monitoring sleep in community-dwelling older adults and PLWD. The low cost of consumer sleep technologies, their ease of deployment, and their remote data monitoring and collection capabilities hold promise for the large-scale implementation of these technologies, including in research settings [6], [7], [8], [9]. These technologies range from devices that detect bed occupancy or wrist activity, to devices that record physiological signals capable of performing epoch-by-epoch sleep staging similar to PSG. Unfortunately, not all consumer sleep technologies, such as wrist-worn trackers and wearable EEG devices, are currently regulated by medical device standards and adhere to American Academy of Sleep Medicine (AASM) sleep scoring guidelines, raising concerns about their accuracy and reliability [10].

The choice of technology deployed for longitudinal monitoring depends on the intended use and the aspect of sleep, behaviour, or condition that is of interest clinically or in a specific research context. The wide variety of wearable and contactless sleep technologies on the market deliver varying levels of accuracy and data quality. The majority of assessments of accuracy and data quality are based on evaluations that are primarily conducted in young and middle-aged adults. Indeed, evaluations of the performance of these devices generally do not include the populations for which they have potential clinical use including older adults or PLWD. A recent systematic review into the use of non-invasive sleep-measuring devices in mid to late life adults did not identify any studies evaluating device accuracy in participants with mild cognitive impairment or Alzheimer’s [11]. To bridge this gap to some extent, there is an increasing number of studies evaluating the promise of consumer sleep technologies in older adult populations and PLWDs [12], [13], [14]. Several wrist wearables and contactless sleep technology devices currently available on the market have been shown to provide acceptable quality in assessing aspects of sleep including timing, continuity, heart rate, and sleep disordered breathing, i.e. sleep apnoea, in older populations and PLWD. However, the accuracy of detection of sleep vs wake quantified by both sensitivity and specificity, and sleep stage classification by these non-EEG devices remains challenging. Wearable devices that employ electroencephalography (EEG) could address these limitations and offer an accurate and user-friendly alternative to portable PSG systems for home sleep assessment [15]. To address this, consumer-grade wearable sleep EEG devices have been developed to provide detailed insights into sleep architecture at home. Current research on sleep and its relation to brain function not only focuses on sleep stages but also on aspects of the sleep EEG, such as slow waves and sleep spindles. Thus, current research questions require that the quality of the EEG signals acquired by these devices is sufficient for quantitative analyses of EEG data. This will contribute to the understanding of sleep’s contribution to disease progression and development of biomarkers [16], [17], [18], [19].

In summary, to realise the potential of consumer grade sleep EEG devices the accuracy of sleep staging and the quality of EEG signals relative to polysomnography needs to be evaluated in relevant populations. Here we provide a comprehensive comparison of Dreem headband (DHB, Dreem, Paris, France [now Beacon Biosignals, Inc, Boston, United States]), a sleep EEG wearable, against PSG in the sleep laboratory in a group of older adults and people living with Alzheimer’s (PLWA). Although several studies have evaluated the DHB in younger adults, to the best of our knowledge there is no published evaluation that compares the DHB both in community-dwelling older adults and PLWA [14], [20], [21], [22]. Here, we compared classical sleep measures and sleep stages provided by the DHB algorithm against polysomnography manually scored by two independent human scorers and evaluated the signal quality and prominent characteristics of the EEG, such as slow-wave activity, sigma activity in non-REM sleep, and rapid eye movements in REM sleep.

## Methods

### Population and Protocol

The participant group consisted of 62 individuals between the ages of 44 and 83 (mean±SD)= 70.5±6.7 years). The Dreem 2 (DHB 2) and 3 (DHB 3) headbands were used to perform an overnight laboratory recording alongside polysomnography in the UK Dementia Research Institute Clinical Research Facility at the Surrey Sleep Research Centre with a 10-hour time-in-bed period. It should be noted that there was a pre-laboratory period involving data collection at home. This first-night in the laboratory with extended time-in-bed protocol was chosen to induce mildly disrupted sleep, which presents a greater computational challenge for algorithms compared to scoring highly consolidated sleep. The data were collected in two separate studies conducted in line with the Declaration of Helsinki and Good Clinical Practice (GCP). The inclusion/exclusion criteria were kept liberal to allow participants with stable co-morbidities to participate if it was safe for them to do so, and the study population to be more similar to the intended use group. Concomitant medications use needed to be stable, and the participants needed to be able to comply with study procedures and perform activities of daily life independently. The population characteristics were also assessed by questionnaires including the Epworth Sleepiness Scale (ESS), Standardized Mini-Mental State Examination (SMMSE), Pittsburgh Sleep Quality Index (PSQI), and International Consultation on Incontinence Questionnaire (ICIQ) (summarised in Table 1). It should be noted that there was a pre-laboratory period involving data collection at home.

**Table 1.** Demographical Characteristics of Participants.

| Characteristics | Study 1 | Study 2 | Total |
| --- | --- | --- | --- |
| **N** | 35 | 27 | 62 |
| **Age (years)**<br>[min max] | 70.8 (4.9)<br>[65 83] | 70.1 (8.7)<br>[44 81] | 70.5 (6.7)<br>[44 83] |
| **Women (n [%])** | 14 [40] | 13 [48] | 27 [44] |
| **Standardized Mini-Mental State Examination (SMMSE)** | 28.7 (1.4)<br>[25, 30] | 28 (1.5)<br>[24, 30] | 28.37 (1.48)<br>[24, 30] |
| **BMI (kg/m<sup>2</sup>)** | 26.7 (4.7)<br>[20 39.7] | 27.6 (6)<br>[16.8 42.1] | 27.1 (5.3)<br>[16.8 42.1] |
| **AHI (events/hr)** | 19.9 (15.5)<br>[1.6 66.7] | 14.8 (11.7)<br>[1.7 48.2] | 17.7 (14.1)<br>[1.6 66.7] |
| **Pittsburgh Sleep Quality Index (PSQI)** | 4.2 (1.9)<br>[1 7] | 4.2 (3.63)<br>[0 14] | 4.1 (2.8)<br>[0 14] |
| **Epworth Sleepiness Scale (ESS)** | 3.7 (2.7)<br>[1 9] | 4.4 (3.3)<br>[0 12] | 4 (2.9)<br>[0 12] |
| **International Consultation on Incontinence Questionnaire (ICIQ)** | 1.0 (1.7)<br>[0 6] | 2.3 (3.2)<br>[0 13] | 1.6 (2.5)<br>[0 13] |
| **PLWA** |  |  |  |
| **N** | - | 12 | 12 |
| **Age (years)**<br>[min max] | - | 73.1 (5.8)<br>[61 81] | 73.1 (5.8)<br>[61 81] |
| **Women (n [%])** | - | 5 [42] | 5 [42] |
| **SMMSE** | - | 26.9 (1.5)<br>[24 29] | 26.9 (1.5)<br>[24 29] |
| **Study Partner** |  |  |  |
| **N** | - | 8 | 8 |
| **Age (years)**<br>[min max] | - | 68.3 (12.9)<br>[44 80] | 68.3 (12.9)<br>[44 80] |
| **Women (n [%])** | - | 5 [63] | 5 [63] |
| **SMMSE** | - | 28.9 (0.8)<br>[28 30] | 28.9 (0.8)<br>[28 30] |
| **Cognitively unimpaired older adults / Controls** |  |  |  |
| **N** | 35 | 7 | 42 |
| **Age (years)**<br>[min max] | 70.8 (4.9)<br>[65 83] | 67 (6.2)<br>[61 79] | 70.2 (5.2)<br>[61 83] |
| **Women (n [%])** | 14 [40] | 3 [43] | 17 [40] |
| **SMMSE** | 28.7 (1.4)<br>[25, 30] | 28.9 (1.1)<br>[27 30] | 28.7 (1.3)<br>[25 30] |

Note: The values shown are the mean followed by the (standard deviation) and [min max].

### Study 1

Study 1 focused on cognitively intact (SMMSE ≥ 27) older adults aged between 65 and 83 years (70.8 ± 4.9 years; N=35; Men: Women = 21: 14) and was conducted using DHB 2 in two cohorts. The Surrey Clinical Research Facility (CRF) recruitment database was used to identify, screen, and recruit participants. The study received a favourable opinion from the University of Surrey Ethics committee (UEC-2019-065-FHMS). A detailed description of the population characteristics and inclusion/exclusion criteria can be found in our description of the protocol [23].

### Study 2

In Study 2, DHB data were collected from 27 participants aged between 44 and 81 years (70.1 ± 8.7 years; N=27; Men: Women = 14: 13). Participants in this study consisted of PLWA (N=12), their caregiver or study partner (N=8), and controls (N=7). Neither the controls and partners were cognitively impaired. The recruitment of cognitively intact older adults was conducted through the Surrey Clinical Research Facility (CRF). PLWA and their respective study partners were identified and recruited through the Surrey and Borders Partnership NHS Foundation Trust (SABP) memory services.

PLWA participating in this study had to have a confirmed diagnosis of prodromal or mild Alzheimer’s disease (AD) and be aged between 50 and 85 years. This included having an SMMSE ≥ 23, residing in the community, and maintaining a stable dose of any dementia medication for a minimum of three months. PLWA had the option to participate either independently or with a designated ‘study partner’. Inclusion criteria for the study partner were that they had known the PLWA for at least six months. They could be a caregiver, family member, or friend, be aged over 18 and have an SMMSE ≥ 27. The inclusion /exclusion criteria for the cognitively intact older adults (aged 50-85 years) were the same as those in Study 1. Study 2 received a favourable opinion from an NHS ethics committee (22/LO/0694) and is registered as a clinical study (ISRCTN10509121).

### Sleep Diary and PSG assessments

In both studies participants maintained a sleep diary. The sleep diary was either paper-based or a digital application based on the consensus sleep diary [24]. PSG data were collected during the in-laboratory session using the SomnoHD system (SOMNOmedics GmbHTM, Germany). The system collected data via electroencephalography (EEG; sampled at 256 Hz from F3-M2, C3-M2, O1-M2, F4-M1, C4-M1, and O2-M1), electrocardiography (sampled at 256 Hz), respiratory inductance plethysmography (RIP) thorax and abdomen (sampled at 128 Hz), photoplethysmography (PPG, sampled at 128 Hz), electromyography (EMG; sampled at 256 Hz, both submental and limb), and electrooculography (EOG; sampled at 256 Hz; E2-M1 and E1-M2). Sleep scoring was performed at 30-second intervals in the DOMINO software environment, following the guidelines of the AASM, by two independent scorers (a Registered Polysomnographic Technologist™ [RPSGT] and a trained scorer), to generate a consensus hypnogram [25]. The AASM recommended scoring rules were applied by the RPGST to determine the apnea-hypopnea index (AHI) and period limb movement index (PLMI) [26].

### Dreem Headband (DHB)

Both DHB 2 and 3 are dry EEG headbands that allow the collection of sleep EEG (sampled at 250 Hz) for multiple days along with 3D accelerometer data (sampled at 50 Hz). The audio stimulation output feature of the devices was disabled in both studies. The firmware and software versions of the DHB used in the studies is provided in Supplemental Table S1.

Both DHB versions (2 and 3) allow the transfer of the collected data to Dreem secure-cloud servers and the data can be downloaded through a common data portal. The automatic sleep stage scoring of the collected data is performed online through Dreem’s proprietary algorithm and available for download in the data portal along with the sleep statistics. Both the sleep stage scoring and sleep statistics are automatically generated by the DHB algorithm for the entire period over which the device is switched on (i.e., over the entire recording period). In both studies, the recordings were manually initiated without the use of the DHB mobile app. The fit of the device to the participant’s head was optimised using the adjustable band provided by the manufacturer to obtain a firm but comfortable device placement. The PSG wire-up was conducted after the DHB was fitted on the head of the participant.

### Dreem 2 Headband (DHB 2)

The DHB 2 comprises of six dry electrodes: four frontal (Fp1, Fp2 [ground], F7 and F8) and two occipital (O1 and O2) electrodes, to acquire the EEG data. The available channels (seven total 7) in the downloaded data are Fp1-O1, Fp1-O2, Fp1-F7, Fp1-F8, F7-O1, F8-O2 and F8-F7. In addition to the EEG and the 3D accelerometer, DHB 2 also used a photoplethysmography (PPG) sensor to collect photoplethysmography data at 50Hz.

### Dreem 3 Headband (DHB 3)

The DHB 2 uses five dry electrodes: three frontal (Fp2 [ground], F7 and F8) and two occipital (O1 and O2) electrodes to collect the EEG data. The available channels (total 5) in the downloaded data are F7-O1, F8-O1, F7-O2, F8-O2, F8-F7.

### Collected data

The DHB data were downloaded from the Dreem data portal. The files consisted of the EEG data (.h5), automatic DHB hypnogram (.txt) and sleep reports (.csv). The DHB scores five stages (Wake, rapid eye movement (REM), N1, N2 and N3) at 30-second intervals along with artifact epochs detected. The sleep report files contain sleep summary measures determined over the recording period of the device and recording quality measures. The PSG data were exported from the DOMINO software interface in European Data Format (.edf) file for the raw EEG and physiological data, and text (.txt) file format for the consensus hypnograms. Two participants (one study partner and one cognitively intact older adult/control) in Study 2 used DHB 2.

### Assessment of Sleep measures

#### Sleep Summary

The sleep summary measures assessed include total sleep time (TST), sleep onset latency (SOL), sleep efficiency (SEFF), wake after sleep onset (WASO), number of awakenings (NAW), and time spent in each sleep stage (N1, N2, N3, rapid eye movement (REM) and non-REM sleep). The PSG sleep summary measures are determined over the lights-off period. The lights-off time was self-selected by the participant and therefore varies between participants. The DHB algorithm automatically generates the sleep summary measures over the entire recording period and is referred to as the analysis period –automatic (AP-A). The primary comparison was performed against the PSG lights-off period and the AP-A measures of the DHB algorithm. Further, to perform a comparison between the PSG and DHB over the same analysis period (AP), we estimated the DHB sleep summary measures over the PSG lights-off period (referred to as the analysis period – manual [AP-M]).

The association between the PSG and DHB were analysed using coefficient of determination, Bland-Altman agreement measures, consistency intraclass correlation with two-way random effects (ICC), standardised absolute differences (SAD) and symmetric mean absolute percentage error (SMAPE). The differences between the compared estimates were tested for normality using Shapiro-Wilk test before Bland-Altman analysis was performed and 95% confidence intervals are provided were applicable [27], [28], [29]. Measures such as minimum detectable change (MDC), bias, and limits of agreement (LoA) were also computed. The MDC represents the smallest detectable change in an estimate that exceeds the device’s measurement error[28]. We relied on the confidence interval range of sleep duration (% TST) associated with interrater variability in scoring reported by Younes et al., to define the satisfactory agreement level for the sleep summary measures (Reference values from Younes et al.,: N1= 11.1±7.1 %TST; N2= 18.9±7.4 %TST; N3= 14.4±6.1 %TST;REM= 10.0±4.7 %TST) [30]. Please note that the DHB sleep report files also contain lights off and lights on variables. These variables are not identical to the lights-off and lights-on times as recorded by the RPSGT technician and used for the PSG scoring. Please refer to the Supplemental Materials, Comparison of the Lights-Off Period section for further discussion of the Lights-off, Lights-On variables as reported in the Dreem sleep. All the data analyses reported here were performed using MATLAB (version 2022b; Math Works).

#### Epoch-by-epoch concordance

The epoch-by-epoch (EBE) concordance of the automatic DHB algorithm hypnogram to the consensus hypnogram was performed over the common recording interval between the PSG and DHB. The measures used to quantify epoch-by-epoch sleep stage prediction performance was determined via standard metrics such as sensitivity, specificity, accuracy, Matthew’s correlation coefficient (MCC) and F1 Score. Both two stage (sleep vs wake) performance and five stage (W, N1, N2, N3, REM) classification performance were evaluated. We relied on the interrater reliability estimates reported in the meta-analysis by Lee et al, to determine the acceptable agreement for EBE concordance [31]. We used MCC rather than kappa due to the identical relationship between the two metrics in the positive quadrant [32].

### Time synchronisation of the DHB and PSG

To perform an accurate comparison of the EEG data collected by the DHB to the gold standard PSG EEG, we time-synchronised the DHB data using synchronization software provided by *‘Dreem Research’* in Study 1. The output of the software consisted of files containing both the synchronised DHB EEG data and synchronised automatically scored DHB hypnogram. For Study 2, the synchronization of the DHB data was performed using an FFT-based synchronization approach described in the Supplemental Materials, DHB Synchronisation section.

### Assessment of similarity of EEG power spectra

The EEG data from both the PSG and DHB were filtered between 0.5 to 35 Hz using a zero-phase filter with third-order Butterworth response (Matlab function ‘*filtfilt*’). To facilitate a robust comparison of the collected EEG data between the PSG and DHB, new PSG EEG channel derivations (F3-O1, F4-O1, F3-O2, F4-O2, F4-F3) were created to match the DHB EEG channels.

### Spectral estimation and Artifact detection

The spectral composition of the EEG channels was determined by segmenting the recorded channels of interest into 30 second epochs using the consensus hypnogram as the temporal marker. Each 30 second epoch was divided into 4 second sub epochs with an overlap of 1 second as described in [33]. Due to the difficulty in securing the device firmly on the participant head and the nature of dry electrode technology used in the DHB, we noticed numerous contact artifacts in the collected DHB EEG signals. Artifacts were determined in the 4 second sub-epochs using the steepness of the spectral slope between 0.75 and 30 Hz as a quality measure [34], [35]. The slope was determined using a first order fit (‘*polyfit’* function). Sub-epochs that had a spectral slope greater than –100 µV/Hz between 0.75 and 30Hz were considered as artifact. We applied this lenient rule which is optimised for sensitivity in detecting artefacts rather than specificity, to both PSG and DHB data. An example DHB 3 recording before and after artifact removal and the average artifact power spectral density (PSD) and Normal EEG PSD for the recording showcasing the method is given in Supplemental Figure S1. It can be seen that the activity from the head mounted accelerometer in DHB is inefficient in determining EEG segments with artifacts and also showcases the effectiveness of the approach used.

For each of the 4 second sub-epochs determined to be artifact-free, PSD estimates with a resolution of 0.25 Hz were computed via Fast Fourier Transform (FFT) after the application of an Hanning window (‘*periodogram’* function). The epoch wise PSD was computed by averaging the PSD estimates of artifact-free 4 second sub-epochs in each epoch (a 30 second epoch is considered valid only if >50% of the data is artifact-free). Following the artifact removal, epoch wise band power estimates were generated. The same estimation procedure was applied to both DHB and PSG data.

The spectral concordance of DHB to PSG was evaluated in two ways: 1. association between the band powers, and 2. Similarity between the PSDs in the 0.5 to 30 Hz range. The five EEG band powers that were assessed included slow wave activity (SWA, 0.75 – 4.5 Hz), Theta activity (4.75 – 7.75 Hz), Alpha activity (8-12 Hz), Sigma activity (11-16 Hz) and Beta activity (15 – 30 Hz) during NREM sleep [36]. Coefficients of determination were determined for each comparison and a one-way repeated measures ANOVA (‘*anova’* function) was performed using a significance level of 0.05 [37].

### Effects of population characteristics on performance

To understand the influence of participant characteristics on the performance of the DHB scoring algorithm, a linear mixed-effects model was fitted to all the sleep measures with participant as the random effect (‘*fitlme’* function). The fixed effects included sex, age, body mass index (BMI), AHI, ESS, ICIQ, PLMI, PSQI, and DHB device type (DHB2 or DHB3). This analysis was performed for two cases, 1. With participant category (PLWA, Caregiver or controls) as fixed effect and 2. Without participant category as fixed effect.

## Results

### Characteristics of the study population

The analysis set (n=62) consisted of 35 men (56%) and 27 (44%) women with a mean age of 70.5 years (SD=6.7). There were 42 controls (mean ± SD= 70.2 ± 5.2 years; Men: Women = 25:17), 12 PLWA (73.1 ± 5.8 years; Men: Women = 7:5) and 8 caregivers (68.3 ± 12.9 years; Men: Women=3:5). Sixty-five percent of the participants (40/62) had one or more self-reported comorbidities including obesity, arthritis, type 2 diabetes, etc. Among the three groups, 92% (11/12) of PLWA, 75% (6/8) of caregivers and 55% (23/42) of controls reported comorbidities. The detailed demographic characteristics of the study population are presented in Table 1. Significant urinary incontinence (n=6; ICIQ>5) was reported by about 10% of the participants. According to self-report, about 75% of the participants did not have any significant sleep disturbances (n=47; PSQI<5) and only 5 % of the participants had significant sleepiness (n=3; ESS>10).

The polysomnography recordings conducted during the laboratory session revealed that 87% (54/62) of the participants had some form of obstructive sleep apnea (AHI>5) with 16% (10/62) having severe apnea (AHI≥30) while 29% (18/62) had a periodic limb movement index >15. The sleep efficiencies of the study population determined by the gold standard laboratory polysomnography during this first night in the laboratory ranged between 25.7% to 92.6% (mean±SD= 67.9±12.7%) with a mean total sleep time of 351 minutes (SD=78.1). Table 2 summarizes the PSG sleep summary measures, and the definitions of the measures are described in the Supplemental Materials.

**Table 2.**
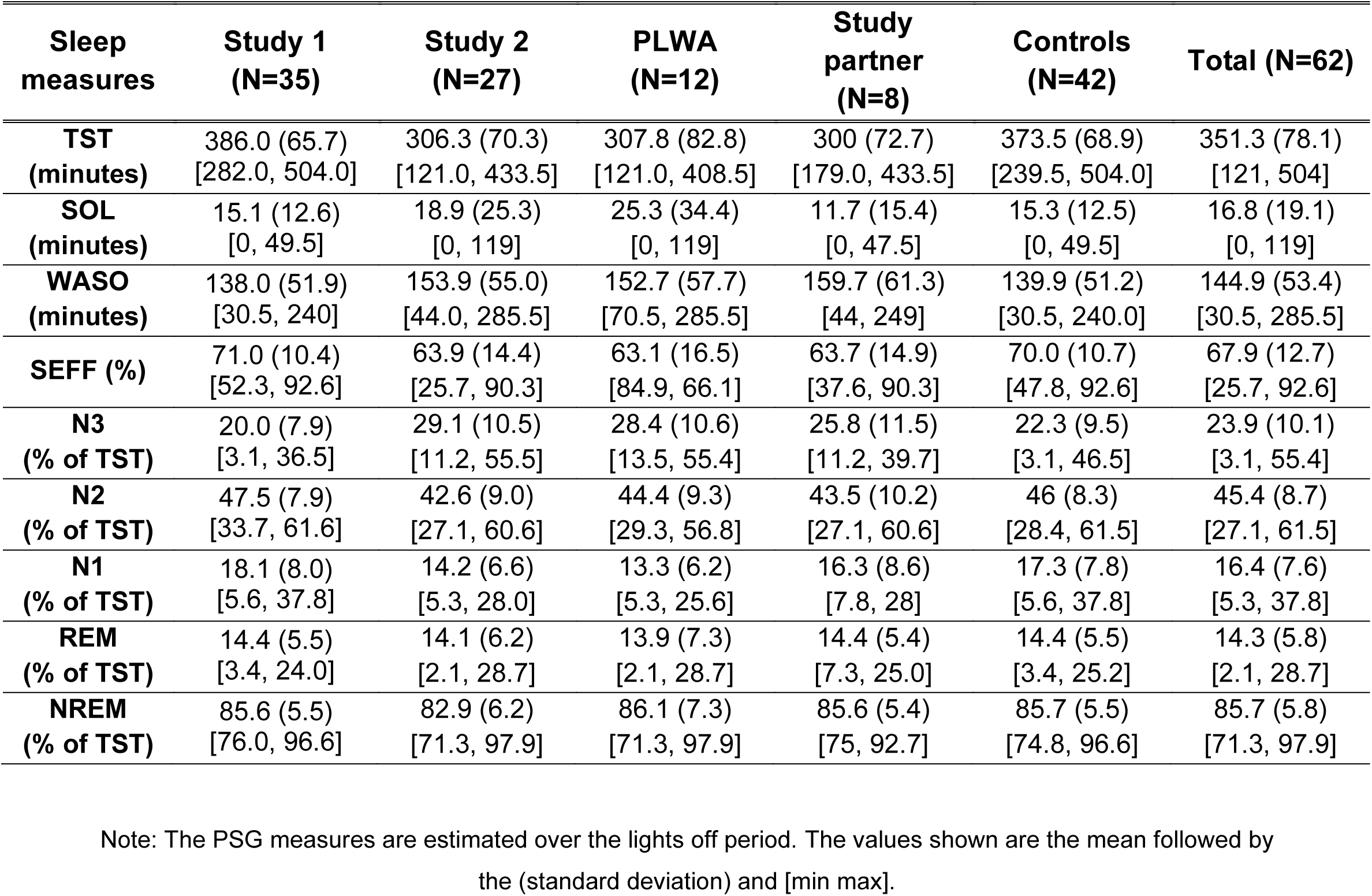
PSG Sleep measures.

| Sleep measures | Study 1 (N=35) | Study 2 (N=27) | PLWA (N=12) | Study partner (N=8) | Controls (N=42) | Total (N=62) |
| --- | --- | --- | --- | --- | --- | --- |
| **TST (minutes)** | 386.0 (65.7)<br>[282.0, 504.0] | 306.3 (70.3)<br>[121.0, 433.5] | 307.8 (82.8)<br>[121.0, 408.5] | 300 (72.7)<br>[179.0, 433.5] | 373.5 (68.9)<br>[239.5, 504.0] | 351.3 (78.1)<br>[121, 504] |
| **SOL (minutes)** | 15.1 (12.6)<br>[0, 49.5] | 18.9 (25.3)<br>[0, 119] | 25.3 (34.4)<br>[0, 119] | 11.7 (15.4)<br>[0, 47.5] | 15.3 (12.5)<br>[0, 49.5] | 16.8 (19.1)<br>[0, 119] |
| **WASO (minutes)** | 138.0 (51.9)<br>[30.5, 240] | 153.9 (55.0)<br>[44.0, 285.5] | 152.7 (57.7)<br>[70.5, 285.5] | 159.7 (61.3)<br>[44, 249] | 139.9 (51.2)<br>[30.5, 240.0] | 144.9 (53.4)<br>[30.5, 285.5] |
| **SEFF (%)** | 71.0 (10.4)<br>[52.3, 92.6] | 63.9 (14.4)<br>[25.7, 90.3] | 63.1 (16.5)<br>[84.9, 66.1] | 63.7 (14.9)<br>[37.6, 90.3] | 70.0 (10.7)<br>[47.8, 92.6] | 67.9 (12.7)<br>[25.7, 92.6] |
| **N3 (% of TST)** | 20.0 (7.9)<br>[3.1, 36.5] | 29.1 (10.5)<br>[11.2, 55.5] | 28.4 (10.6)<br>[13.5, 55.4] | 25.8 (11.5)<br>[11.2, 39.7] | 22.3 (9.5)<br>[3.1, 46.5] | 23.9 (10.1)<br>[3.1, 55.4] |
| **N2 (% of TST)** | 47.5 (7.9)<br>[33.7, 61.6] | 42.6 (9.0)<br>[27.1, 60.6] | 44.4 (9.3)<br>[29.3, 56.8] | 43.5 (10.2)<br>[27.1, 60.6] | 46 (8.3)<br>[28.4, 61.5] | 45.4 (8.7)<br>[27.1, 61.5] |
| **N1 (% of TST)** | 18.1 (8.0)<br>[5.6, 37.8] | 14.2 (6.6)<br>[5.3, 28.0] | 13.3 (6.2)<br>[5.3, 25.6] | 16.3 (8.6)<br>[7.8, 28] | 17.3 (7.8)<br>[5.6, 37.8] | 16.4 (7.6)<br>[5.3, 37.8] |
| **REM (% of TST)** | 14.4 (5.5)<br>[3.4, 24.0] | 14.1 (6.2)<br>[2.1, 28.7] | 13.9 (7.3)<br>[2.1, 28.7] | 14.4 (5.4)<br>[7.3, 25.0] | 14.4 (5.5)<br>[3.4, 25.2] | 14.3 (5.8)<br>[2.1, 28.7] |
| **NREM (% of TST)** | 85.6 (5.5)<br>[76.0, 96.6] | 82.9 (6.2)<br>[71.3, 97.9] | 86.1 (7.3)<br>[71.3, 97.9] | 85.6 (5.4)<br>[75, 92.7] | 85.7 (5.5)<br>[74.8, 96.6] | 85.7 (5.8)<br>[71.3, 97.9] |

Note: The PSG measures are estimated over the lights off period. The values shown are the mean followed by the (standard deviation) and [min max].

### Example laboratory recording

An example of overnight data collected during the laboratory session is depicted in Figure 1. The subfigures consist of the hypnogram, the spectrogram and the slow wave activity during sleep of a PSG channel (F3O2) and a DHB channel (F7O2). The comparison of the hypnogram reveals that although there is overall good structural similarity between the hypnogram, the DHB is less sensitive in detecting short bouts of wake (underestimates WASO by 18.5 minutes) and is biased towards scoring N2 (overestimates N2 by 77.5 minutes).

**Figure 1.**
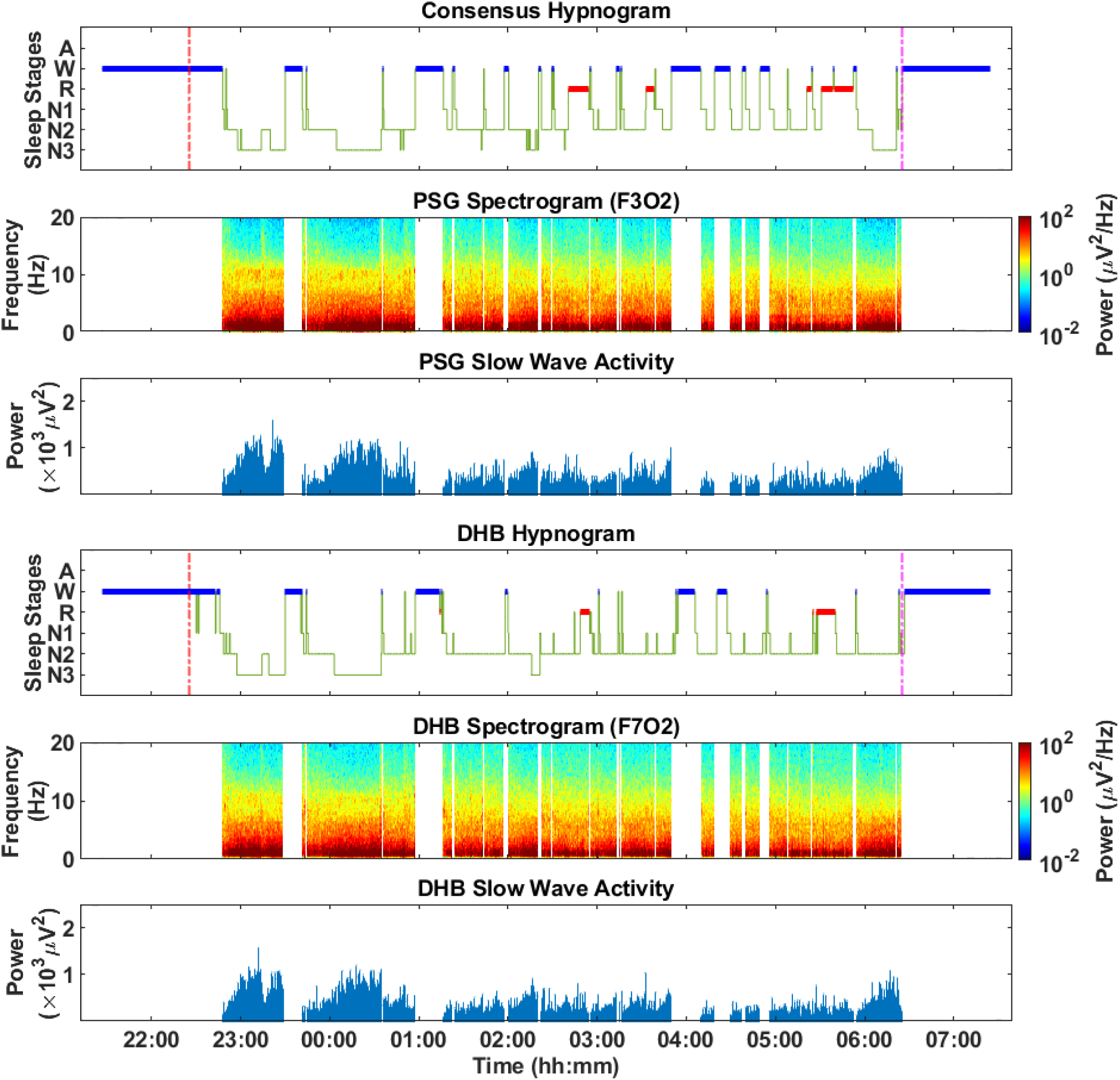
Data collected in the laboratory from a male PLWA participant between the ages of 70 and 75. The consensus polysomnography (PSG) hypnogram is depicted at the top of the plot followed by the PSG spectrogram, slow wave activity (SWA), Dreem headband (DHB) hypnogram, SWA and spectrogram. Epochs scored as Wake have been removed to showcase the SWA profile during sleep.

The PSG and DHB spectrograms after artifact removal are shown in Figure 1. The signal quality and similarity of EEG collected by the DHB can be inferred from inspection of the spectrograms and the associated SWA plots. The frequency distribution and power trends in both spectrogram and SWA plot between PSG and DHB closely follow each other, with DHB having a larger number of artifacts (eg., spikes in the power due to electrode contact changes) and relatively lower power in the higher frequencies. The number of 4 second sub-epochs classified as artifact in DHB data (7% of TST) was also high compared to PSG (4% of TST).

The associations between the various raw spectral band powers for each 30 second epoch are depicted in the scatter plot shown in Supplemental Figure S2. The plots show high association (r^2^=0.64, p<0.001) between PSG and DHB SWA and a low to moderate level of association (r^2^=0.24 to 0.44, p<0.001) for rest of the spectral bands.

### Assessment of sleep measures

#### Sleep summary measures

The sleep summary assessment of the automatic DHB algorithm estimates (analysis period automatically determined by the device [AP-A]) is presented in Table 3 and 4 and Figures 2 and 3. The DHB algorithm estimates (AP-A) of TST and SEFF were accurate with a SMAPE of less than 10% and high agreement (r^2^>0.65; p<0.001) with acceptable estimation bias. WASO was underestimated with moderate agreement while SOL was overestimated (See Table 3). The total number of awakening (NAW, awakenings ≥1min) determined by the DHB also had a moderate agreement with PSG. The ICC between DHB and PSG was the highest for TST (0.90) followed by SEFF (0.83), WASO (0.61) and SOL (0.59).

**Figure 2.**
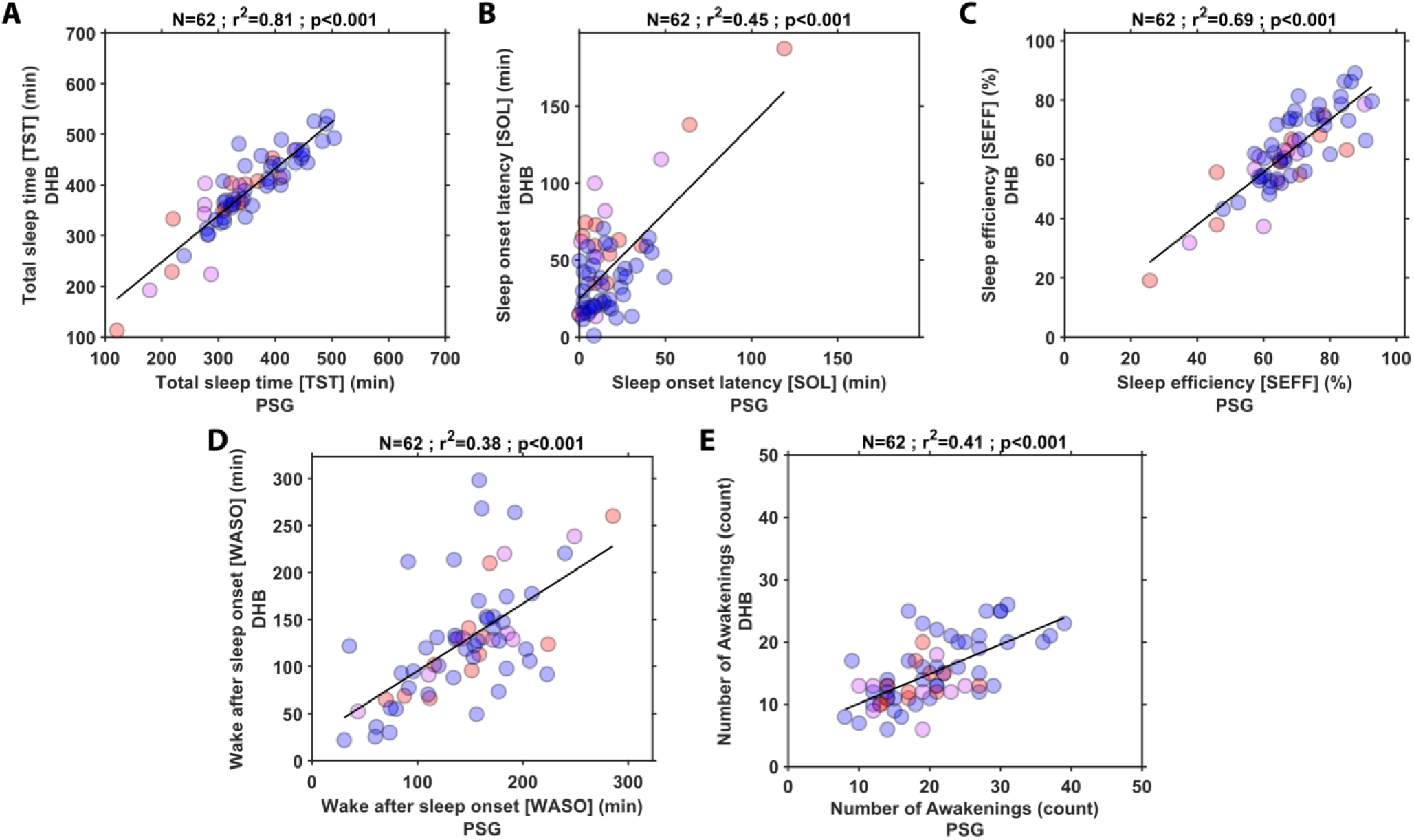
Association between polysomnography (PSG) and Dreem headband (DHB) for total sleep time (TST), sleep onset latency (SOL), sleep efficiency (SEFF), wake after sleep onset (WASO) and number of awakenings. The data points in red depicts people living with Alzheimer’s, magenta depicts caregivers, and blue depicts controls. The DHB measures are automatically estimated by the DHB algorithm over the recording period [AP-A]. The top of each of the plots shows the number of participants, the coefficient of determination and significance value of the association between the devices.

**Figure 3.**
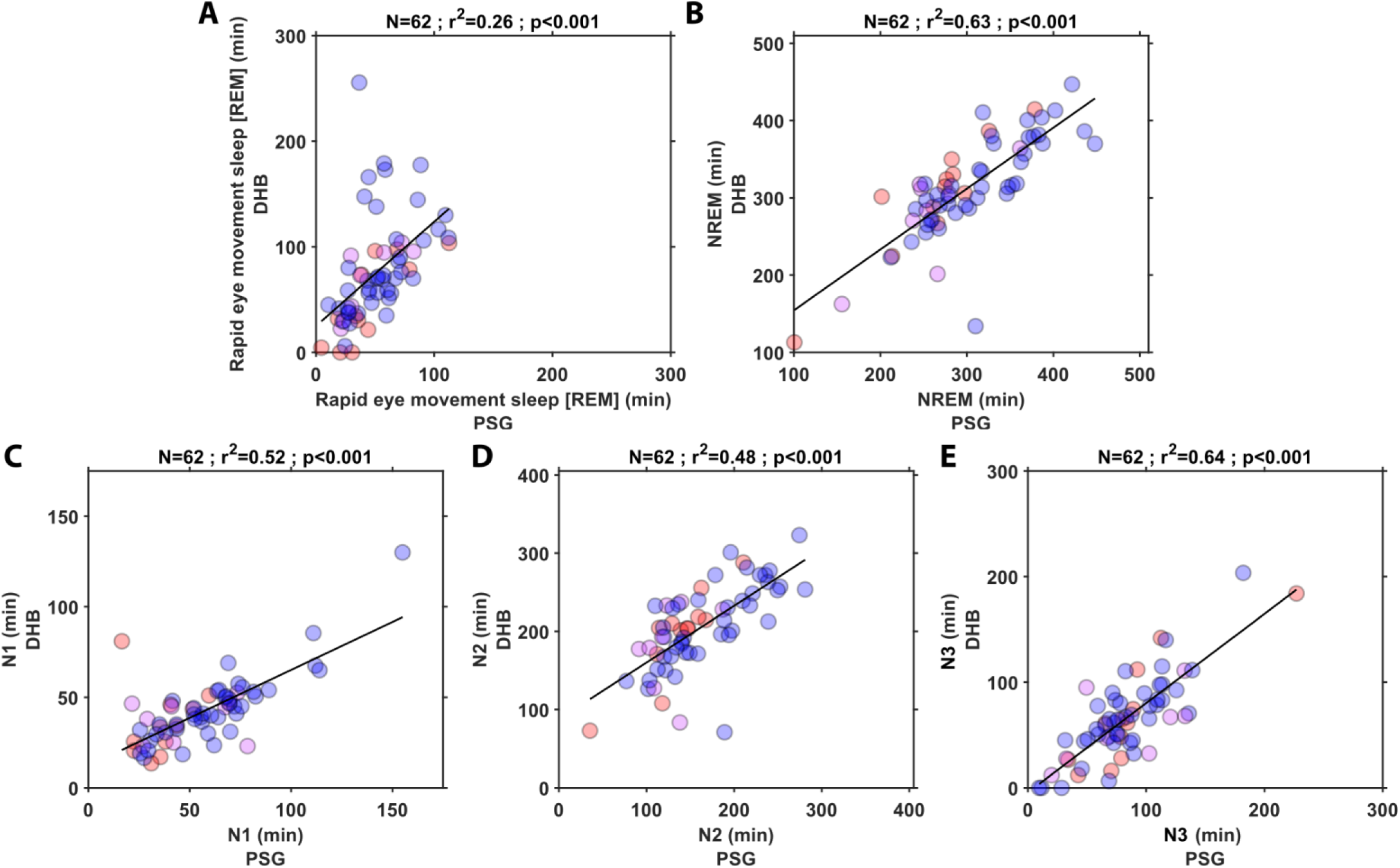
Association between polysomnography (PSG) and Dreem headband (DHB) for rapid eye movement (REM), non REM (NREM), N1, N2 and N3 sleep durations. The data points in red depicts people living with Alzheimer’s, magenta depicts caregivers, and blue depicts controls. The DHB measures are automatically estimated by the DHB algorithm over the recording period [AP-A]. The top of each of the plots shows the number of participants, the coefficient of determination and significance value of the association between the devices.

**Table 3.** Agreement Metrics for All-Night Sleep Summary Measures (AP-A)

| **Sleep measure** | **DHB Mean (SD)** | **PSG Mean (SD)** | **Bias [95% CI]** | **LoA Lower bound [95% CI]** | **LoA Upper bound [95% CI]** | **MDC** | **SAD [95% CI]** | **SMAPE [95% CI]** | **ICC [95% CI]** |
| --- | --- | --- | --- | --- | --- | --- | --- | --- | --- |
| **SOL (min)** | 43.9 (32.2) | 16.9 (19.1) | 27 (23.9) [20.9, 33.1] | -20 [-30.4, -9.5] | 73.9 [63.5, 84.4] | 46.9 | 1.1 [1.1, 1.3] | 51.5 [51.5, 58.8] | 0.59 [0.4, 0.73] |
| **TST (min)** | 387.7 (80) | 351.4 (78) | 36.3 (35.6) [27.3, 45.4] | -33.5 [-49.1, -18] | 106.2 [90.6, 121.7] | 69.8 | 0.5 [0.5, 0.8] | 5.6 [5.6, 6.8] | 0.9 [0.84, 0.94] |
| **WASO (min)** | 127.5 (62.3) | 144.7 (53.5) | -17.2 (51.5) [-30.3, -4.1] | -118.2 [-140.7, -95.7] | 83.9 [61.4, 106.3] | 101 | 0.7 [0.7, 1] | 16.1 [16.1, 19.5] | 0.61 [0.42, 0.74] |
| **SEFF (%)** | 62.7 (13.5) | 67.9 (12.7) | -5.2 (7.6) [-7.2, -3.3] | -20.2 [-23.5, -16.9] | 9.7 [6.4, 13] | 14.9 | 0.6 [0.6, 0.8] | 6 [6, 7.1] | 0.83 [0.73, 0.89] |
| **NAW (counts)** | 30.4 (10.8) | 39.7 (16.4) | -9.3 (11.8) [-12.3 -6.3] | -32.5 [-37.7 -27.4] | 13.9 [8.7 19.1] | 23.2 | 0.9 [0.9 1.1] | 16.4 [16.4 19.3] | 0.63 [0.46 0.76] |

Note. Number of participants, N=62. AP-A- sleep measures automatically generated by the DHB algorithm over the DHB recording period. The values shown are the mean followed by the (standard deviation) and [95% confidence interval]. The metrics include Bias – difference in measurement between the Dream headband (DHB) and PSG; Lower and Upper bounds of the Bias; Minimum detectable change (MDC) – smallest detectable change independent of measurement error (half of Bland Altman agreement width); Standardized absolute difference (SAD) – directionless version of Cohen's d; Symmetric mean absolute percentage error (SMAPE) – mean error in measurement expressed as percentage; Absolute intraclass correlation with two-way random effects (ICC) – measures of measurement reliability; The ICC estimates were computed for each participant and the mean, and 95 % confidence interval is reported. All the values are rounded to two decimal places.

For the sleep stage duration estimations (AP-A, see Table 3), the highest agreement with PSG was observed for N3 sleep, even though it was underestimated (-17.2 minutes). For REM sleep, DHB overestimated its duration, and the agreement was low (24.4 minutes; SMAPE = 23.6%; r^2^=0.26; p<0.001). N1 was underestimated (-14.4 minutes) and N2 duration was overestimated (43.5 minutes) and for both these sleep stages the agreement with PSG was moderate (see Figure 3)

When the sleep summary measures for the DHB were computed for the PSG lights-off period (AP-M), rather than the period automatically detected by the DHB, the concordance between PSG and DHB improved compared to the automatic DHB estimates (AP-A). The sleep stage duration performance over the lights off period (AP-M) for N3 and REM followed that of the DHB algorithm estimates (AP-A) while the accuracy of the N1 and N2 estimates decreased. The SOL estimate on the other hand was more accurate over the lights off period (AP-M). The results of the lights off analysis are presented in Supplemental Table S2, S3 and Figure S3.

When expressed as % TST, the automatic DHB sleep duration (AP-A, see Table 4) estimates yielded high agreement with PSG measures for NREM (N1 and N3) sleep (r^2^>0.4; p<0.001) and low agreement with N2 and REM sleep (r^2^≤0.15; p<0.01) while the estimates over the lights off period (AP-M) had lower agreement for both NREM and REM (r^2^≤0.3) (see Table 4, and Supplemental Table S3 and Figure S4).

**Table 4.**
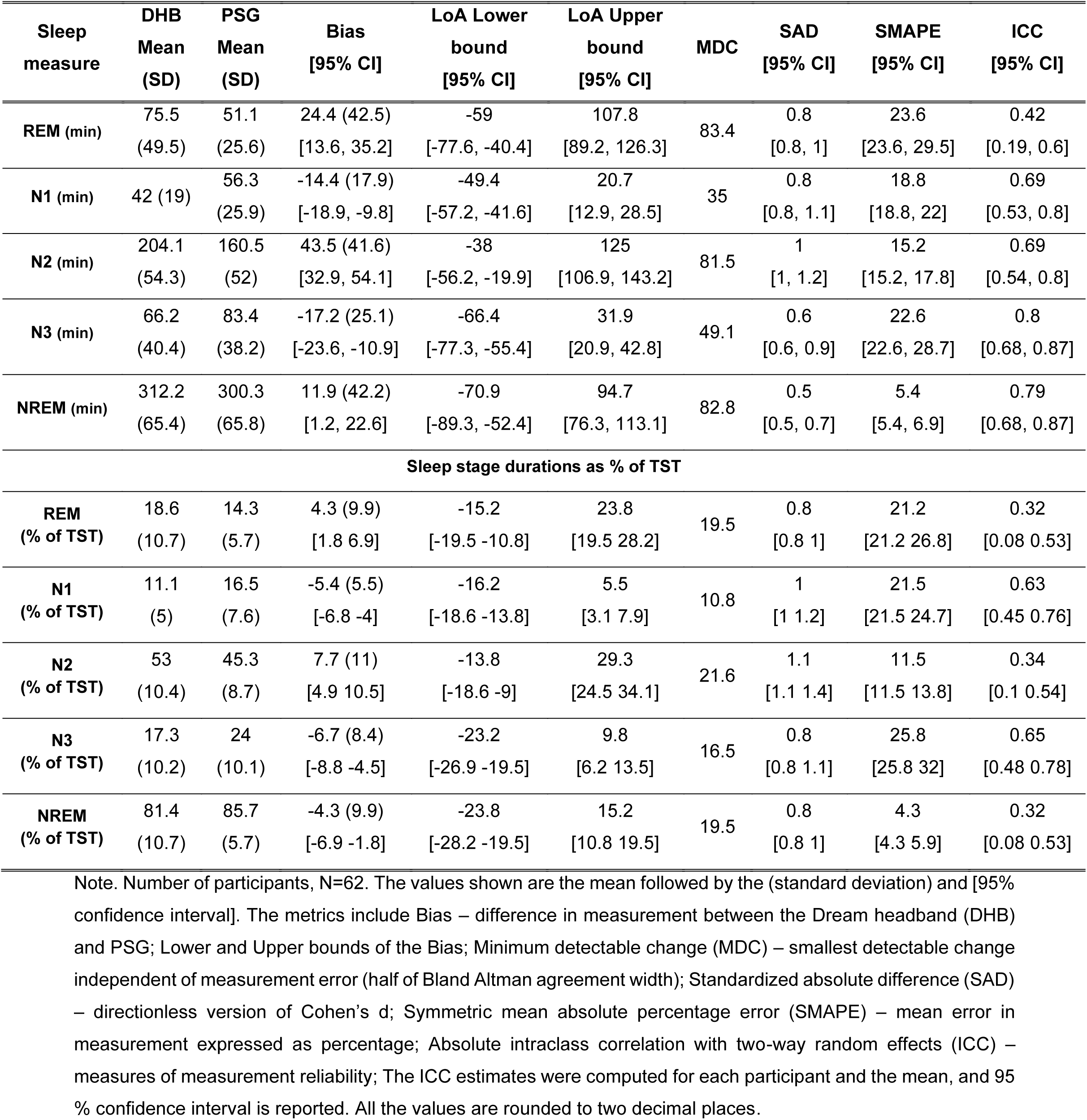
Agreement Metrics for Sleep duration measures (AP-A)

When the linear mixed effects (LME) model was fit with participant as the random effect for the difference between the in-laboratory DHB measures and the PSG measures without the participant category information, there were significant effects of device (i.e., DHB3 compared to DHB 2) on SOL (overestimation of 23.8 minutes by DHB2, p=0.0013), duration and % TST of REM (underestimation of -37.88 minutes by DHB2, p=0.008 and -8.1%, p=0.011), N2 (overestimation of 29.5 minutes, p=0.029 and 10.15%, p=0.003 by DHB2) and NREM (overestimation of 30.2 minutes, p=0.025 and 8.1%, p=0.011 by DHB2). We also found significant effects (p<0.05) of AHI on N2, NREM and REM sleep. There were no significant effects of MMSE or participant category.

#### Epoch-by-epoch (EBE) concordance

The DHB had a high discriminative power to distinguish between sleep and wake with a MCC of 0.77. Among the sleep stages, the NREM and N3 sleep stages were most accurately classified (MCC >0.65) followed by REM and N2 (see Table 5). DHB performed the poorest in classifying N1 sleep (MCC=0.18). The detailed EBE concordance metrics are provided in Table 5 and the pooled confusion matrix is shown in Figure 4.

**Figure 4.**
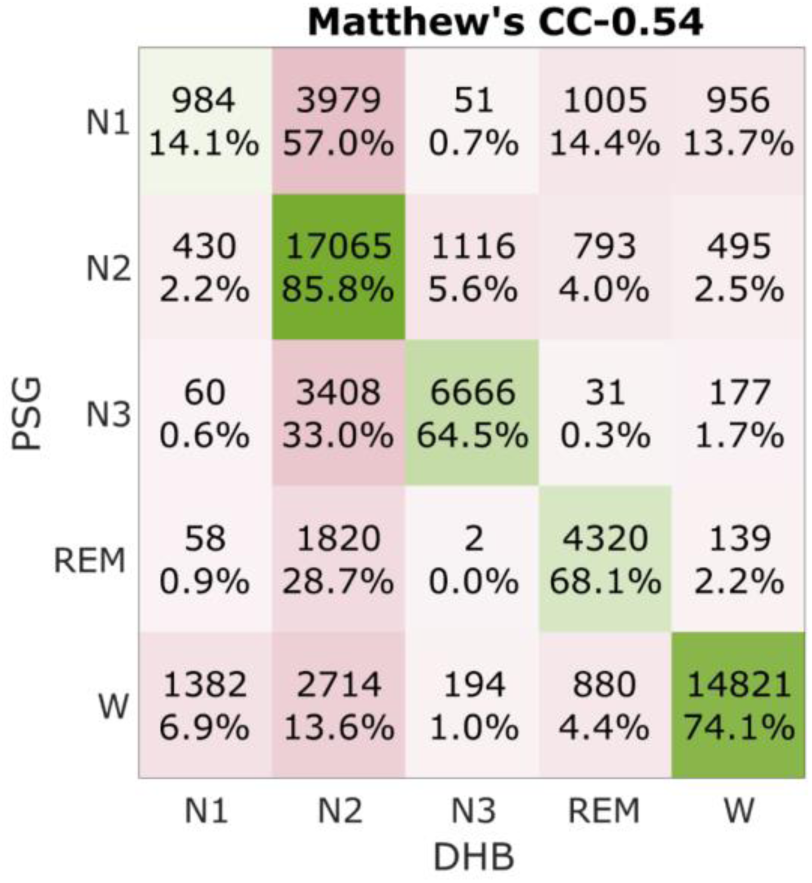
Confusion Matrix for DHB sleep stage classification: Epoch-by-epoch concordance. The pooled confusion matrix is for all 62 participants. The total number of epochs were 63546 over the common recording period.

**Table 5.**
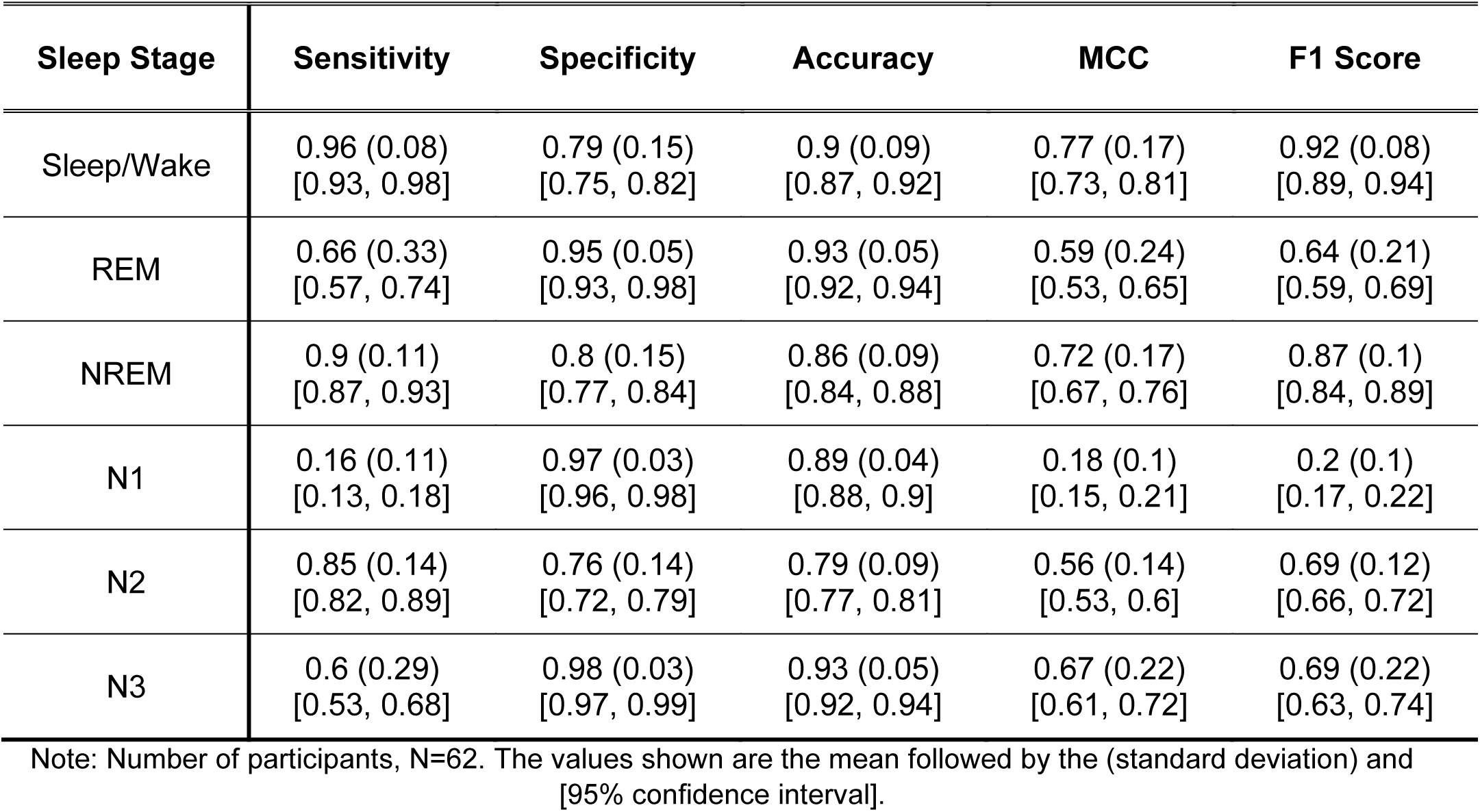
Epoch-by-epoch concordance over the common recording interval.

| **Sleep Stage** | **Sensitivity** | **Specificity** | **Accuracy** | **MCC** | **F1 Score** |
| --- | --- | --- | --- | --- | --- |
| Sleep/Wake | 0.96 (0.08)<br>[0.93, 0.98] | 0.79 (0.15)<br>[0.75, 0.82] | 0.9 (0.09)<br>[0.87, 0.92] | 0.77 (0.17)<br>[0.73, 0.81] | 0.92 (0.08)<br>[0.89, 0.94] |
| REM | 0.66 (0.33)<br>[0.57, 0.74] | 0.95 (0.05)<br>[0.93, 0.98] | 0.93 (0.05)<br>[0.92, 0.94] | 0.59 (0.24)<br>[0.53, 0.65] | 0.64 (0.21)<br>[0.59, 0.69] |
| NREM | 0.9 (0.11)<br>[0.87, 0.93] | 0.8 (0.15)<br>[0.77, 0.84] | 0.86 (0.09)<br>[0.84, 0.88] | 0.72 (0.17)<br>[0.67, 0.76] | 0.87 (0.1)<br>[0.84, 0.89] |
| N1 | 0.16 (0.11)<br>[0.13, 0.18] | 0.97 (0.03)<br>[0.96, 0.98] | 0.89 (0.04)<br>[0.88, 0.9] | 0.18 (0.1)<br>[0.15, 0.21] | 0.2 (0.1)<br>[0.17, 0.22] |
| N2 | 0.85 (0.14)<br>[0.82, 0.89] | 0.76 (0.14)<br>[0.72, 0.79] | 0.79 (0.09)<br>[0.77, 0.81] | 0.56 (0.14)<br>[0.53, 0.6] | 0.69 (0.12)<br>[0.66, 0.72] |
| N3 | 0.6 (0.29)<br>[0.53, 0.68] | 0.98 (0.03)<br>[0.97, 0.99] | 0.93 (0.05)<br>[0.92, 0.94] | 0.67 (0.22)<br>[0.61, 0.72] | 0.69 (0.22)<br>[0.63, 0.74] |

Note: Number of participants, N=62. The values shown are the mean followed by the (standard deviation) and [95% confidence interval].

The DHB predicted N1 and N3 more often as N2, which contributed to the lower performance in distinguishing each NREM stage separately even though performance for overall NREM classification remained good. The EBE concordance, as measured by Matthew’s correlation coefficient also showed a significant effect of DHB3 (LME model: an increase of 0.11 in MCC, p=0.014) compared to DHB 2.

### Assessment of similarity of EEG power spectra

To investigate the similarity of EEG spectra of PSG and DHB signals, DHB recordings were selected if, for at least one EEG channel, 90% of the 30-second epochs contained >50% artifact-free 4-second sub-epochs. After applying these criteria, 73% (45/62) of DHB recordings in the laboratory were deemed usable for the spectral analysis (mean±SD= 602±141 epochs). For all the analysis reported below, the PSG EEG channel derivation that was similar to the selected low artifact DHB channel was used.

Normalised band powers were computed and averaged across all NREM epochs in each recording (here, each 4 second sub-epochs was normalised using the power within 0 and 35 Hz as 100%). Figure 5 illustrates the association between the different spectral band power estimates from PSG and DHB. SWA, theta and alpha activity estimates of PSG and DHB were found to be significantly correlated (r^2^>0.56; p<0.001). The association was found to be moderate (r^2^=0.34; p<0.001) for the sigma band and poor (r^2^=0.08; p=0.058) for the beta band. Further inspection of the lack of association revealed a significant difference (p<0.01, one-way ANOVA) in the slopes of the correlation between PSG and DHB estimates of the average nightly NREM band powers estimated among the participants in Cohort 1 of Study 1 (N=12) and the remaining participants (N=33) in the data. ANOVA revealed that the difference was associated with variations in the firmware versions of the Dreem 2 headband (see Supplemental Table S1).

**Figure 5.**
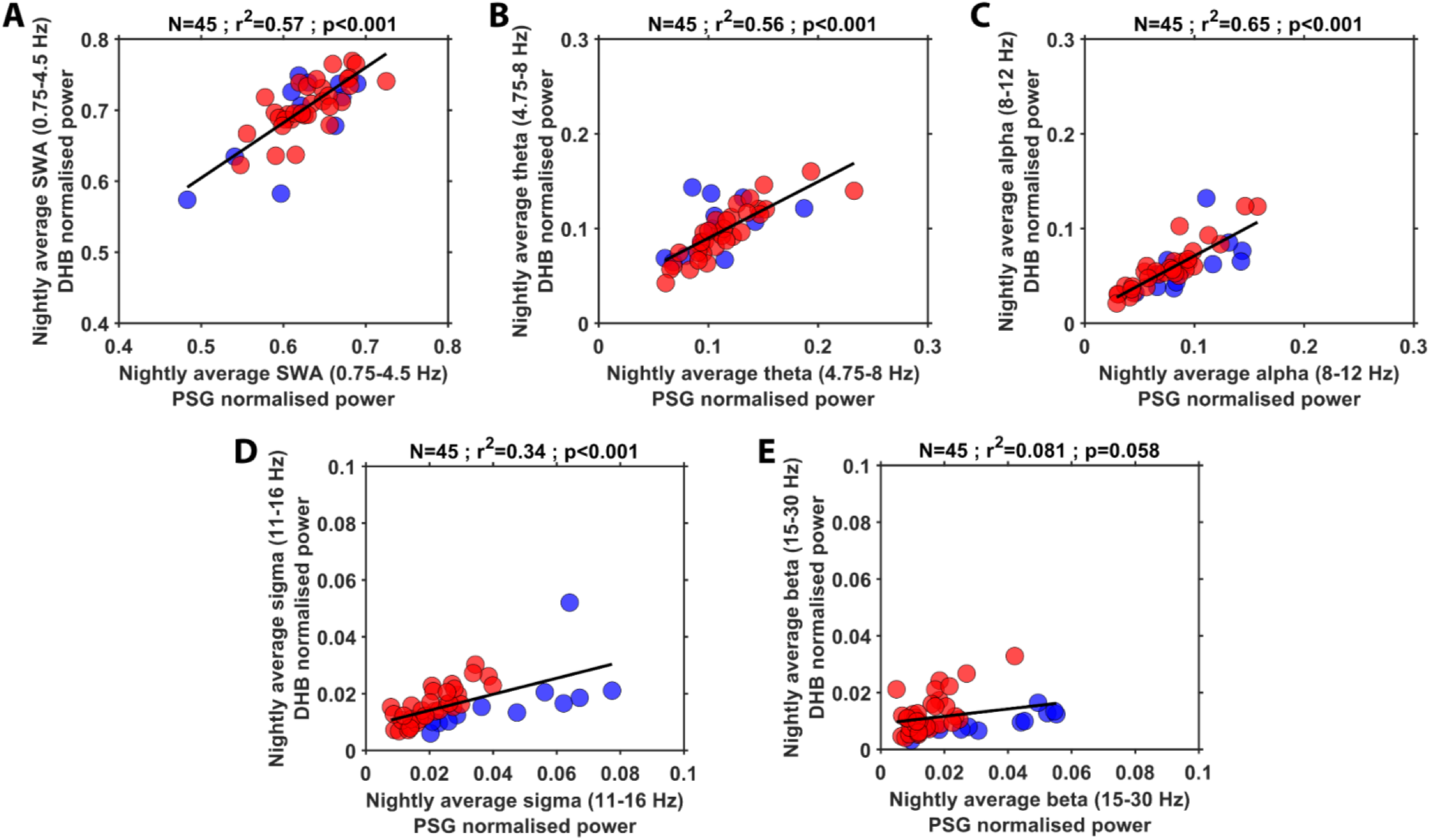
Spectral band power correlation between PSG and DHB recordings. The data points in red depicts data collected in cohort 2 of study 1 and entire study 2, and blue depicts cohort 1 of study 1. The data points are coloured to distinguish the differences in firmware between the two sets of participants. The top of each of the plots shows the number of participants, the coefficient of determination and significance value of the association between the devices.

To assess the similarity between the DHB and PSG PSDs, log PSDs of PSG were compared to log PSDs from DHB data for the EEG channel with the least artifacts, where mutual pairs of 4-second sub-epochs were available. The average Pearson correlation (significance p<0.05), across all participants was (n=444373): ρ =0.61±0.16 [min:0.18, max:0.93]; wake (n=143457) ρ =0.49±0.15 [min:0.18, max:0.93]; sleep (n=300916) ρ =0.66±0.12 [min:0.18, max:0.92]. The ANOVA revealed that the term ‘DHB device type’ had a significant effect on ρ, which were overall higher for DHB 3 compared to DHB 2 (F_(1,444369)_ = 11.48, p <0.001). We have provided an averaged PSD across all participants for all artifact-free epochs in DHB and PSG in the supplemental materials, Figure S5.

## Discussion

The comparison of the Dreem headband (DHB), and associated sleep scoring algorithm, to gold standard PSG measures of sleep, manually scored, and the sleep EEG revealed both strengths and limitations of this wearable sleep EEG device when used in a heterogeneous group of 62 older adults including PLWA.

PSG recordings confirmed prevalent sleep disorders within this population, including obstructive sleep apnea (87% of the participants) and periodic limb movements (29%), which are consistent with the prevalence previously reported in community-dwelling older adults [38], [39]. The 10-hour time-in-bed protocol implemented along with these sleep disorders induced mildly disrupted sleep with an average sleep efficiency of 68% and varying TST (351.3±78.1 minutes) and WASO (144.9±53.4 minutes). Despite this, the mean PSG estimated sleep stage durations (%TST) in our population were in line with the age specific normal ranges reported in the literature [40].

### Challenges in DHB Recording

All laboratory recordings were successful, and DHB data were collected alongside PSG for all 62 participants. The DHB algorithm scored sleep stages for all 62 recordings. However, the quality of the EEG signal from the DHB system was found to be affected by the contact quality of the DHB’s dry electrode with the participant’s scalp. Despite the DHB being used concurrently with PSG and its wear being supervised to ensure optimal placement, design issues specific to the DHB (such as slipping out of position and tilting during the night) and the limitations of the dry electrode contributed to suboptimal signal quality in several recordings. As a result, only 73% (45/62) of the recordings in this study were usable for Quantitative EEG analysis. This issue was further compounded by the lack of referential single channels in the acquired data. Consequently, even minor head movements in active sleepers caused shifts in device positioning, leading to the loss of multiple EEG channel derivations, which is suboptimal. Our observations are consistent with similar issues of optimal device wear, data loss and signal quality issues of the DHB dry electrode system previously discussed in studies of younger populations [41], [42], [43].

### Analysis Period of DHB

The period over which the sleep measures are estimated influences their clinical usefulness and impacts the comparability of the wearable derived sleep measure. The DHB algorithm automatically generates the sleep statistics based on the entire recording period i.e. the period over which the device was switched on, while in standard PSG sleep statistics are estimated over the lights off period. In our analysis, this difference in the analysis period between the DHB algorithm and PSG affected both SOL and WASO estimate accuracy. We found that the accuracy of the SOL improved significantly when the AP was set to the PSG experimental lights off periods which highlights the importance of measuring environmental light to understand the attempted sleep period.

### DHB Algorithm’s Accuracy in Older Adults: A Comparison to Literature

The minimum detectable change (MDC) for sleep duration measures in N1 (MDC = 10.8% TST), N2 (MDC = 21.6% TST), and N3 (MDC = 16.5% TST) stages was within the upper bounds of interscorer variability differences reported in the meta-analysis by Younes et al., 2018 [30]. This suggests that the DHB algorithm performs comparably to manual scoring variability for NREM sleep stages. In contrast, the MDC for REM sleep (MDC = 19.5% TST) exceeds the variability limits outlined in the Younes et al. study, highlighting a need for improvements in detecting REM sleep.

In this study, conducted on older adults (N=62 and mean age= 70.5 ± 6.7 years), the DHB algorithm demonstrated robust performance in epoch-by-epoch sleep/wake classification of sleep/wake with an MCC of 0.77±0.17. Since MCC and kappa are identical to each other we directly compare MCC and kappa in the rest of this discussion [32]. This performance was comparable to the prior evaluations in younger populations, including DHB 3 evaluated by Ong et al., 2023 (N = 40, mean age = 38.03 ± 14.74 years, κ = 0.76 ± 0.12) and DHB 2 evaluated by Arnal et al., 2021 (N = 25, mean age = 35.32 ± 7.51 years, κ = 0.74 ± 0.15) [14], [20]. For specific sleep stages, the DHB algorithm demonstrated moderate accuracy (0.35 < MCC < 0.7) for NREM, N2, N3, and REM stages, while achieving notably lower accuracy for N1 (MCC = 0.18). These results for NREM stages are consistent with values reported by Lee et al., 2022 [31]. Our findings showcase the limitations of the DHB algorithm’s in detecting N1 and REM sleep and suggest that this limitation is likely due to the absence of additional physiological signals such as electrooculography (EOG) and electromyography (EMG).

Overall, the DHB algorithm achieved moderate concordance across all vigilance states (Wake, N1, N2, N3 and REM) with the consensus PSG hypnogram (MCC = 0.55 ± 0.14) in this population of older adults. This performance was lower than that reported in younger populations, where Ong et al., 2023 (Overall κ = [0.76, 0.86]) and Arnal et al., 2021 (Overall κ = 0.74 ± 0.10) demonstrated higher accuracy [14], [20]. This discrepancy is likely attributable to the influence of age-related changes in sleep physiology affecting the DHB algorithm’s performance. Please note that comparison with previous studies is hampered by the limited number of performance metrics reported in those previous studies.

### Comparison of DHB to Other Sleep Monitoring Technologies

To understand the position of the DHB as a sleep monitoring technology in the current landscape of consumer sleep trackers, we compared the performance of the DHB, a dry EEG based wearable, in our older adult population to consumer sleep trackers including wristworn trackers, radars and undermattress trackers evaluated in recent studies. In the study by Ong et al., 2023, actigraphy and consumer sleep trackers (Oura ring and Fitbit) had a lower sleep/wake classification performance compared to DHB 3 (κ = 0.76 ± 0.12, Ong et al., 2023), with the Oura ring (κ = 0.61±0.15) performing better than Fitbit (κ = 0.55±0.14) and Actigraph (κ = 0.46±0.15), showcasing DHB’s superior sleep/wake classification performance due to its direct measurement of brain activity [27], [44], [45]. Comparing DHB performance to our previous evaluation of contactless sleep technologies in older adults (N=35, mean age = 70.8 ± 4.9 years), the Somnofy radar, which provided the higher sleep/wake classification (MCC=0.63±0.12) compared to under-mattress trackers like Withings sleep analyser (MCC=0.41±0.15) and Emfit QS (MCC=0.35±0.16), still under performed relative to DHB [26]. Thus, overall DHB offers a clear advantage over traditional actigraphy, wearable, and contactless sleep trackers with respect to performance. The other devices may however be more suitable for longitudinal recordings. Furthermore, similar to other dry-EEG based sleep technologies like the novel in-ear/ behind-the-ear EEG sensors, DHB faces difficulty in detecting N1 and REM sleep when compared to gold standard PSG [46], [47], [48].

### Suitability of DHB for Quantitative EEG

Despite the use of lenient artifact removal criteria, over 25% of the laboratory recordings were unusable due to high levels of EEG artifacts, highlighting challenges in collecting high-quality EEG signals in wearable dry-EEG devices, even in controlled laboratory settings. Among the remaining 73% of usable recordings, strong correlations were observed for SWA, theta, and alpha band power estimates between PSG and DHB recordings (r² > 0.56; p < 0.001), and moderate correlations were found for the sigma band (r² = 0.34; p < 0.001), suggesting that the DHB reliably captures key sleep EEG features. This offers opportunities to better quantify the associations between sleep and physiological and cognitive outcome measures in larger scale sleep studies in the community.

## Conclusion

This evaluation of the DHB in a sleep laboratory setting assessed its performance in an older population living with health and sleep conditions common in this age group. The concordance between gold standard PSG and DHB algorithm scoring of sleep/wake epochs was substantial (MCC: 0.77), surpassing the interrater agreement (κ=0.70) reported in Lee et al., 2022 [31]. While the accuracy of determining N3 and total NREM sleep durations was moderate, the DHB’s automated algorithm requires further refinement to improve N1 and REM sleep detection and better adapt to the unique characteristics of older adult sleep patterns. Despite the need for improvements in the dry-electrode technology used in DHB, 73% of the recordings were of sufficient quality to quantify slow wave and sleep spindle activity. However, agreement between PSG and DHB estimates of spectral power deteriorated at higher EEG frequencies. Overall, our comprehensive evaluation highlights the DHB’s strengths in terms of portability andperformance relative to other currently available sleep technologies and underscores its potential to become a useful tool for objective monitoring of sleep in community dwelling older adult populations.

## Acknowledgement

This work was primarily supported by the UK Dementia Research Institute, Care Research & Technology Centre at Imperial College, London and the University of Surrey, Guildford, United Kingdom [core award numbers UKDRI-7005, CF2023\7 and UKDRI-7206] which receives its funding from UK DRI Ltd, funded by the UK Medical Research Council, Alzheimer’s Society and Alzheimer’s Research UK. In part supported by NIHR Oxford Health Biomedical Research Centre [NIHR 203316]. The authors thank the staff members Damion Lambert, Marta Messina Pineda and Tegan Ward and the members of the Clinical Research Facility for their help with data collection and curation. They would also like to extend their thanks to the members of Surrey and Borders Partnership (SABP) for help with the recruitment and screening of participants.

KR conducted the data exploration and analysis and prepared the manuscript. DJD conceived the studies and contributed to writing of the manuscript. CdM, GA, HH, RN and VR contributed to the design of the studies, and were responsible for participant recruitment and screening, and study conduct. CdM and GA scored the PSG records and created the consensus hypnogram. The devices were setup and deployed by KR and datasets were downloaded and curated by KR and GA. All the authors contributed to finalising the manuscript.

## Conflicts of interest

DJD received equipment from Somnofy, and is a consultant to Boehringer-Ingelheim and Danisco Sweeteners OY. The other authors declare no competing financial or non-financial interests.

## Data Availability

The data used in this study are available from the co-author Ciro della Monica upon reasonable request. Contact.

## Notes

### Clinical Protocols

https://doi.org/10.3390/clockssleep6010010

### Author Declarations

Ethics committee of University of Surrey (UEC-2019-065-FHMS) and an NHS ethics committee (22/LO/0694) gave ethical approval for this work. This study is also registered as a clinical study (ISRCTN10509121)

