## Supplemental Materials for "Evaluation of Dreem headband for sleep staging and EEG spectral analysis in people living with Alzheimer’s and older adults"

Name: Dr Kiran Kumar Guruswamy Ravindran

Address: Surrey Sleep Research Centre, University of Surrey, GU2 7XP

**Table S1 Firmware and software versions used in the study**

| Dreem Headband | Total participants | Firmware version | Software version |
|---|---|---|---|
| DHB 2 Cohort 1 | 18 | 4.0.85+OCTAVEPROTOCOL1 | 1.121.2 to 1.125.0 |
| DHB 2 Cohort 2 | 17 | 4.0.85+OCTAVEPROTOCOL1 | 1.134.3 to 1.134.4 |
| DHB 3 | 27 | 5.7.15+PRODUCTION | 0.2.30 |

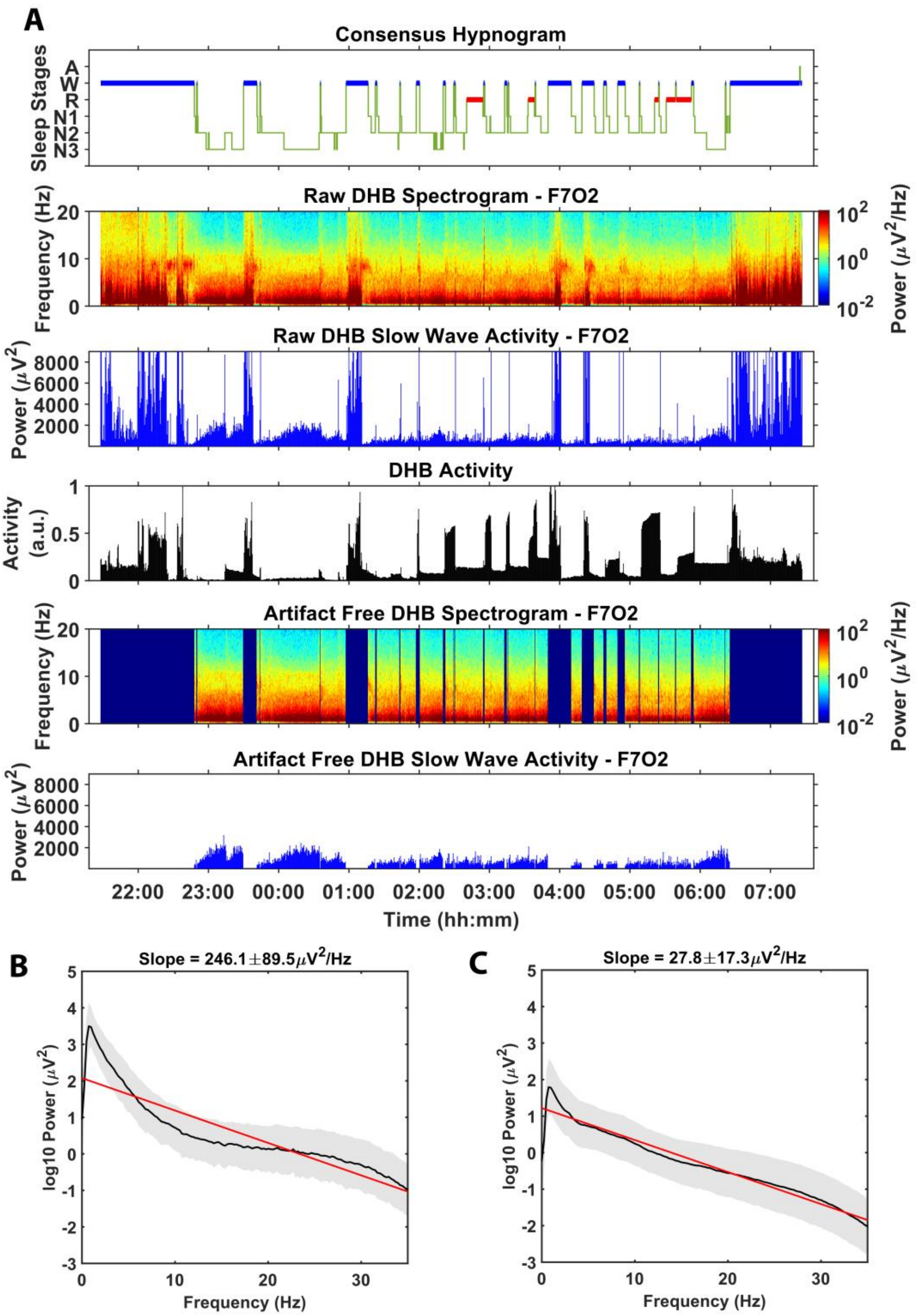


**Figure S1. An example DHB 3 recording to demonstrate the impact of artifact removal. A.** Spectrogram and Slow wave activity before (top) and after artifact removal**.** Epochs scored as Wake have been removed to showcase the slow wave activity (SWA) profile during sleep. **B** average power spectral density (PSD) of artifacts **C.** average PSD of artefact free EEG PSD. Artifacts were determined in the 4 second sub-epochs using the steepness of the spectral slope between 0.75 and 30 Hz as a quality measure. The slope was determined using a first order fit (‘polyfit’ function). Sub-epochs that had a spectral slope greater than –100 µV/Hz between 0.75 and 30Hz were considered as artifact. The 4s sub-epochs for artifacts (853 sub-epochs) and normal EEG (11087 sub-epochs) for the overnight recording are pooled and the average (solid line) and standard error (shaded area) are depicted here. The red line depicts first order fit with the slope depicted on top of the plots.

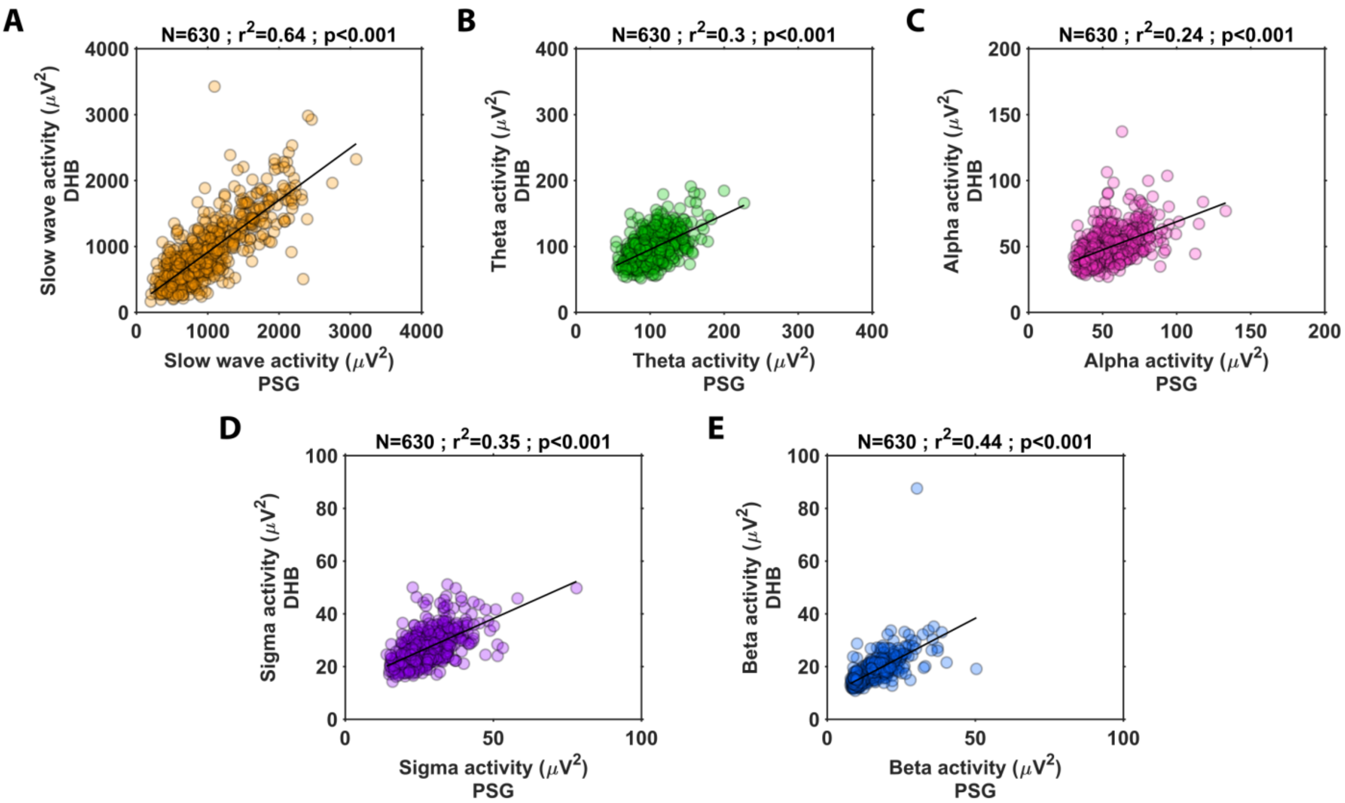


**Figure S2. Raw spectral band powers of the PLWA participant depicted in Figure 1**. A. Slow wave activity (SWA, 0.75 – 4.5 Hz), B. Theta activity (4.75 – 7.75 Hz), C. Alpha activity (8-12 Hz), D. Sigma activity (11-16 Hz) and E. Beta activity (15 – 30 Hz). Only the sleep epochs as determined by polysomnography (PSG) containing >60 % artifact free 4s segments were used. The total number of epochs, the coefficient of determination and the significance value of the association between the DHB and PSG measures are depicted at the top of the scatter plots.

**Table S2. All-night sleep summary measure agreement metrics for all participants calculated over Lights-off period (AP-M)**

| Sleep measure | DHB Mean (SD) | PSG Mean (SD) | Bias [95% CI] | LoA Lower bound [95% CI] | LoA Upper bound [95% CI] | MDC | SAD [95% CI] | SMAPE [95% CI] | ICC [95% CI] |
|---|---|---|---|---|---|---|---|---|---|
| **SOL (min)** | 16.6 (20.6) | 16.9 (19.1) | -0.3 (9.1) [-2.6 2.1] | -18.1 [-22.1 -14.1] | 17.6 [13.6 21.6] | 17.8 | 0.3 [0.3 0.5] | 19.9 [19.9 26.4] | 0.89 [0.83 0.94] |
| **TST (min)** | 379.8 (83.2) | 351.4 (78) | 28.3 (34.9) [19.5 37.2] | -40.1 [-55.3 -24.9] | 96.8 [81.5 112] | 68.4 | 0.4 [0.4 0.7] | 4.7 [4.7 5.8] | 0.91 [0.85 0.94] |
| **WASO (min)** | 118.6 (54.3) | 144.7 (53.5) | -26 (32.3) [-34.2 -17.8] | -89.3 [-103.4 -75.2] | 37.2 [23.1 51.3] | 63.2 | 0.6 [0.6 0.8] | 13 [13 15.7] | 0.82 [0.72 0.89] |
| **SEFF (%)** | 73.4 (13.6) | 67.9 (12.7) | 5.5 (6.6) [3.8 7.2] | -7.5 [-10.3 -4.6] | 18.5 [15.6 21.4] | 13 | 0.5 [0.5 0.7] | 4.7 [4.7 5.8] | 0.87 [0.8 0.92] |

Sleep summary measures calculated over Lights-off period (i.e. Analysis Period-Manual) Note. The values shown are the mean followed by the (standard deviation) and [95% confidence interval]. The metrics include Bias – difference in measurement between the Dream headband (DHB) and PSG; Lower and Upper bounds of the Bias; Minimum detectable change (MDC) – smallest detectable change independent of measurement error (half of Bland Altman agreement width); Standardized absolute difference (SAD) – directionless version of Cohen’s d; Symmetric mean absolute percentage error (SMAPE) – mean error in measurement expressed as percentage; Absolute intraclass correlation with two-way random effects (ICC) – measures of measurement reliability; The ICC estimates were computed for each participant and the mean, and 95 % confidence interval is reported. All the values are rounded to two decimal places.

**Table S3. Sleep duration measures calculated over Lights-off period (AP-M)**

| Sleep measure | DHB Mean (SD) | PSG Mean (SD) | Bias [95% CI] | LoA Lower bound [95% CI] | LoA Upper bound [95% CI] | MDC | SAD [95% CI] | SMAPE [95% CI] | ICC [95% CI] |
|---|---|---|---|---|---|---|---|---|---|
| **REM (min)** | 56.8 (36.1) | 51.1 (25.6) | 5.7 (32.3) [-2.5 13.9] | -57.7 [-71.8 -43.6] | 69 [54.9 83.1] | 63.3 | 0.8 [0.8 1] | 26.3 [26.3 32.5] | 0.47 [0.25 0.64] |
| **N1 (min)** | 23.6 (14.6) | 56.3 (25.9) | -32.8 (29.9) [-40.3 -25.2] | -91.3 [-104.3 -78.3] | 25.8 [12.7 38.8] | 58.5 | 1.7 [1.7 2] | 44 [44 50.8] | - |
| **N2 (min)** | 234.5 (87.8) | 160.5 (52) | 74 (67.4) [56.9 91.1] | -58.2 [-87.6 -28.7] | 206.1 [176.7 235.5] | 132.1 | 1.1 [1.1 1.4] | 19.5 [19.5 22.3] | 0.56 [0.37 0.71] |
| **N3 (min)** | 64.9 (41.7) | 83.4 (38.2) | -18.5 (30.5) [-26.3 -10.8] | -78.3 [-91.6 -65] | 41.2 [27.9 54.5] | 59.7 | 0.7 [0.7 1] | 24.9 [24.9 31.3] | 0.71 [0.56 0.81] |
| **NREM (min)** | 323 (80.5) | 300.3 (65.8) | 22.7 (49.7) [10 35.3] | -74.8 [-96.5 -53.1] | 120.2 [98.5 141.9] | 97.5 | 0.5 [0.5 0.8] | 6.2 [6.2 7.6] | 0.77 [0.65 0.86] |
| **Sleep stage durations as % of TST** | | | | | | | | | |
| **REM (% of TST)** | 14.9 (9.1) | 14.3 (5.7) | 0.6 (8.8) [-1.6 2.9] | -16.6 [-20.4 -12.8] | 17.9 [14 21.7] | 17.2 | 0.8 [0.8 1.1] | 26.1 [26.1 32.4] | 0.33 [0.09 0.54] |
| **N1 (% of TST)** | 6.8 (4.8) | 16.5 (7.6) | -9.7 (8.3) [-11.8 -7.6] | -25.9 [-29.5 -22.3] | 6.5 [2.9 10.1] | 16.2 | 1.7 [1.7 1.9] | 45.7 [45.7 52.4] | 0.15 [0 0.38] |
| **N2 (% of TST)** | 60.8 (14.2) | 45.3 (8.7) | 15.6 (13.8) [12.1 19.1] | -11.4 [-17.4 -5.4] | 42.5 [36.5 48.6] | 27 | 1.5 [1.5 1.7] | 15.6 [15.6 18.1] | 0.32 [0.08 0.52] |
| **N3 (% of TST)** | 17.5 (10.9) | 24 (10.1) | -6.5 (10) [-9 -3.9] | -26.2 [-30.6 -21.8] | 13.2 [8.8 17.6] | 19.7 | 0.9 [0.9 1.2] | 27.7 [27.7 34.3] | 0.54 [0.34 0.7] |

Sleep summary measures calculated over Lights-off period (i.e. Analysis Period-Manual) Note. The values shown are the mean followed by the (standard deviation) and [95% confidence interval]. The metrics include Bias – difference in measurement between the Dream headband (DHB) and PSG; Lower and Upper bounds of the Bias; Minimum detectable change (MDC) – smallest detectable change independent of measurement error (half of Bland Altman agreement width); Standardized absolute difference (SAD) – directionless version of Cohen's d; Symmetric mean absolute percentage error (SMAPE) – mean error in measurement expressed as percentage; Absolute intraclass correlation with two-way random effects (ICC) – measures of measurement reliability; The ICC estimates were computed for each participant and the mean, and 95 % confidence interval is reported. All the values are rounded to two decimal places.

**Table S4. Epoch by Epoch concordance over the lights off period[AP-M]**

| Sleep Stage | Sensitivity | Specificity | Accuracy | Matthew's CC | F1 Score |
|---|---|---|---|---|---|
| Sleep/Wake | 0.95 (0.09) [0.93 0.98] | 0.73 (0.16) [0.68 0.77] | 0.89 (0.09) [0.87 0.91] | 0.73 (0.17) [0.69 0.77] | 0.92 (0.08) [0.90 0.94] |
| REM | 0.66 (0.33) [0.57 0.74] | 0.95 (0.05) [0.94 0.96] | 0.93 (0.05) [0.87 0.91] | 0.59 (0.24) [0.53 0.66] | 0.64 (0.21) [0.59 0.70] |
| NREM | 0.9 (0.11) [0.87 0.93] | 0.76 (0.16) [0.72 0.8] | 0.85 (0.09) [0.83 0.87] | 0.68 (0.17) [0.64 0.73] | 0.87 (0.09) [0.85 0.89] |
| N1 | 0.16 (0.1) [0.13 0.18] | 0.97 (0.03) [0.96 0.97] | 0.88 (0.04) [0.86 0.89] | 0.17 (0.1) [0.15 0.2] | 0.2 (0.1) [0.17 0.22] |
| N2 | 0.85 (0.14) [0.81 0.88] | 0.73 (0.15) [0.69 0.77] | 0.77 (0.09) [0.75 0.79] | 0.54 (0.14) [0.51 0.58] | 0.69 (0.12) [0.66 0.72] |
| N3 | 0.61 (0.28) [0.54 0.68] | 0.97 (0.04) [0.96 0.98] | 0.92 (0.05) [0.91 0.93] | 0.67 (0.21) [0.61 0.72] | 0.69 (0.22) [0.63 0.75] |

Note: AP-M - Analysis Period-Manual i.e., the PSG lights off period. Number of participants, N=62.
The values shown are the mean followed by the (standard deviation) and [95% confidence interval].

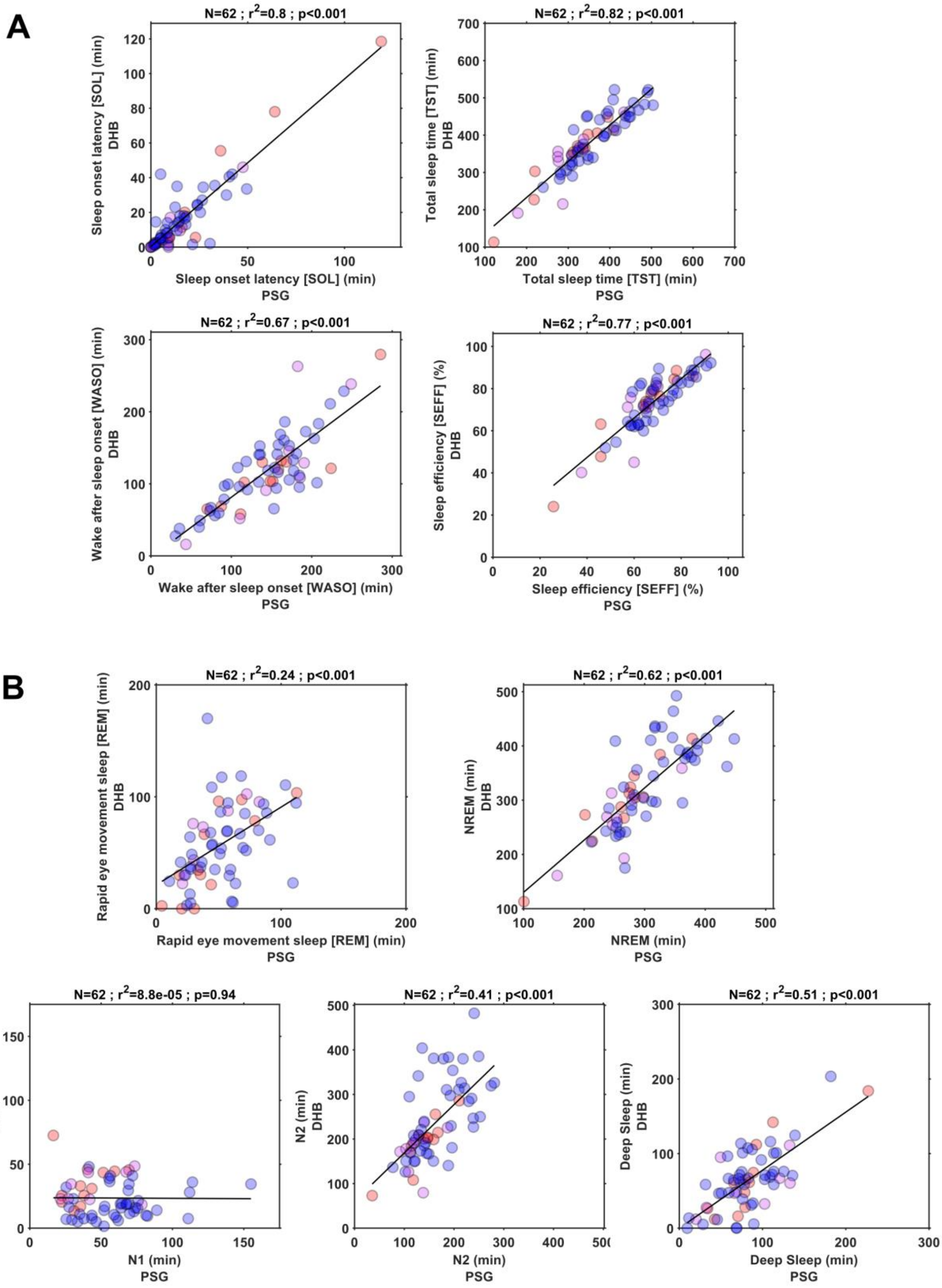


**Figure S3. Scatter plots depicting the association between polysomnography (PSG) and Dreem headband (DHB) for a. Sleep measures and b. Sleep stage duration measures.** The measures are computed over the lights off period of the PSG. The data points in red depicts people living with Alzheimer's, magenta depicts caregivers, and blue depicts healthy older adults. The top of each of the plots shows the number of participants, the coefficient of determination and significance value of the association between the devices.

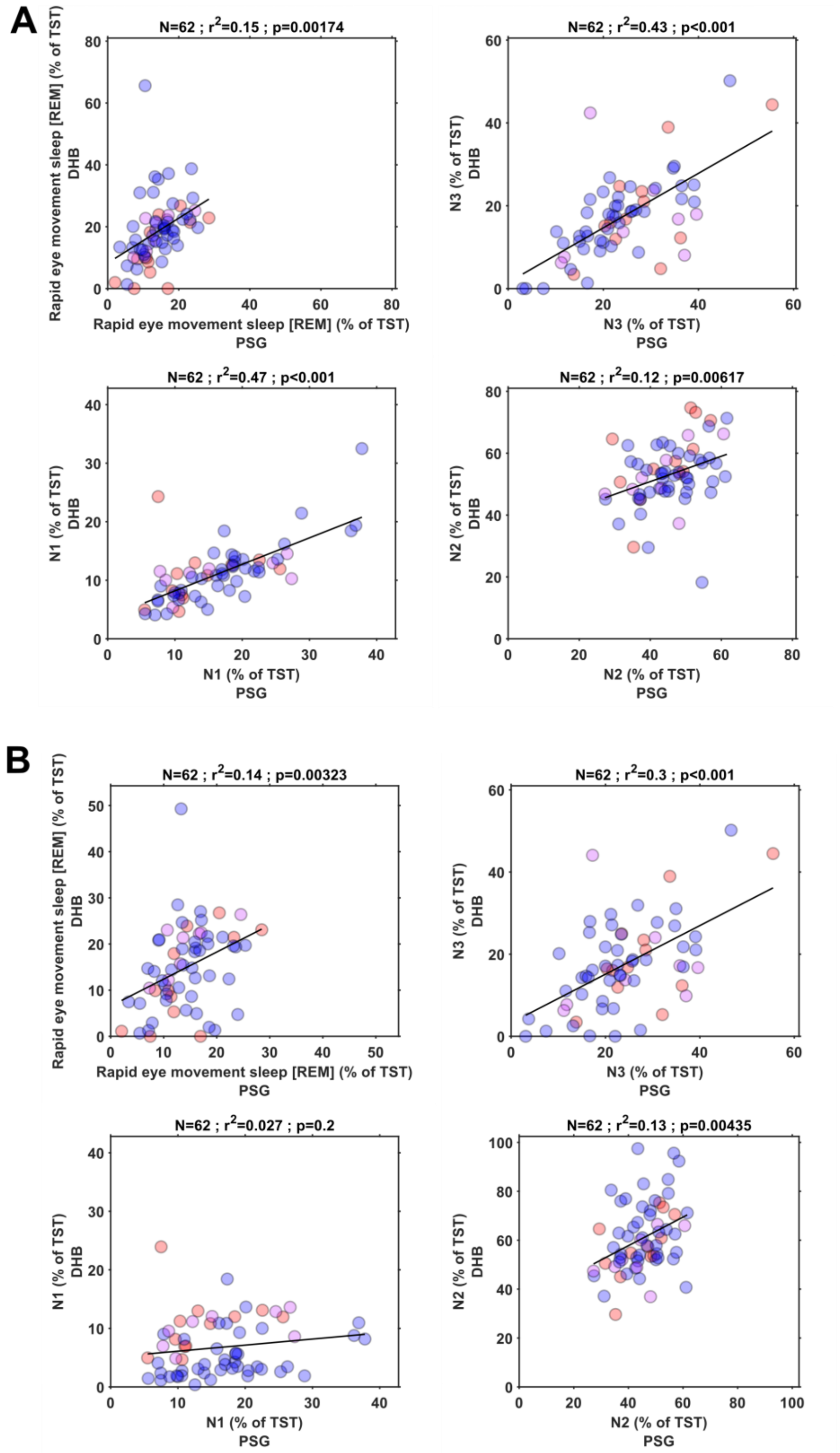


**Figure S4. Scatter plots depicting the association between the sleep stage durations (%total sleep time) of polysomnography (PSG) and Dreem headband (DHB) for a. Automatic DHB measures and b. Measures computed over the lights off period of PSG.** The measures are computed over the lights off period of the PSG. The data points in red depicts people living with Alzheimer's, magenta depicts caregivers, and blue depicts healthy older adults. The number of participants, the coefficient of determination and significance value of the association between the devices is given at the top of each plot.

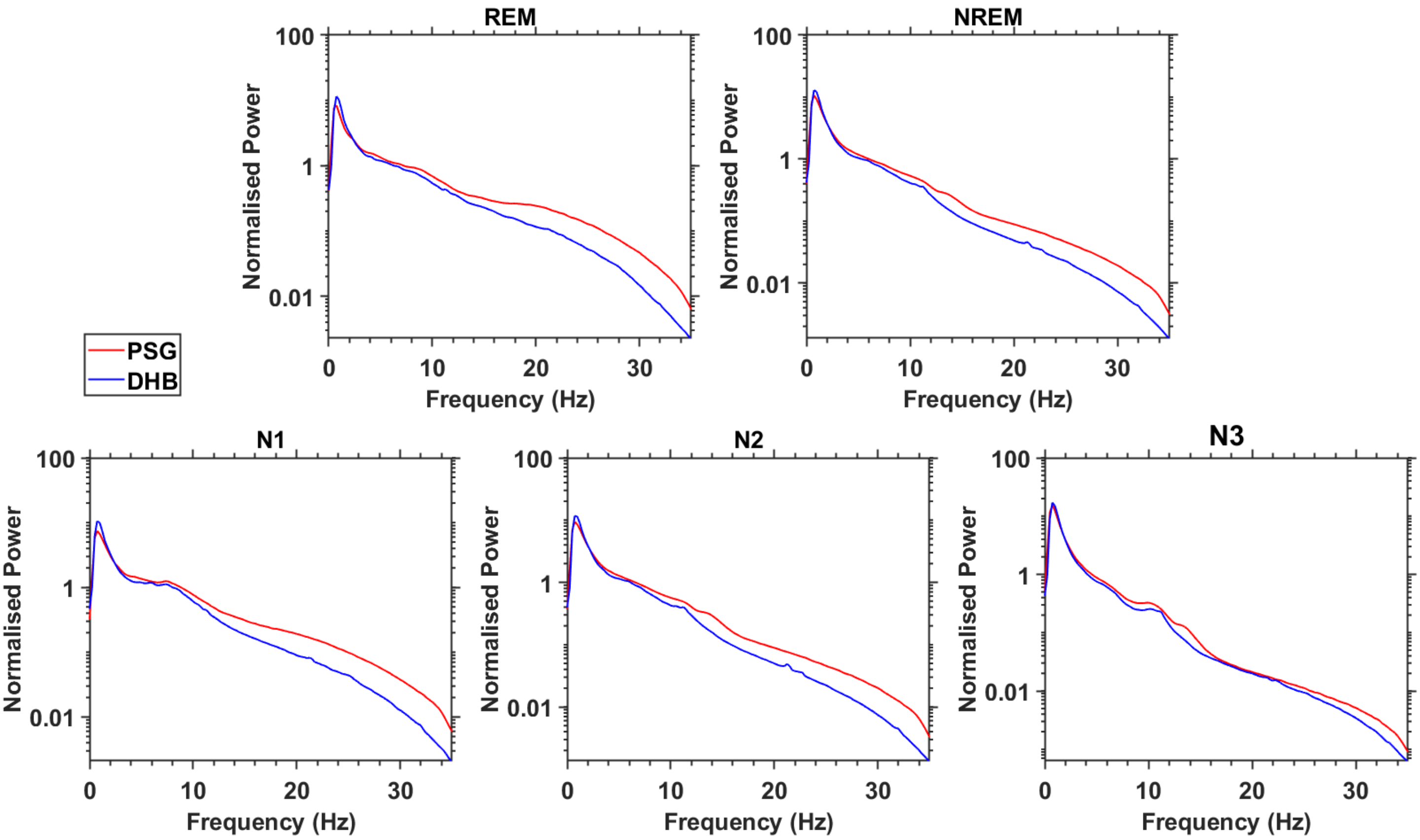


**Figure S5. Average normalised power spectral density (PSD) (in log scale) for the different vigilance states across all participants with contributing data (N=45).** The solid line shows the mean.

### Dreem Headband (DHB) Synchronisation

Although polysomnography (PSG) and DHB data were recorded simultaneously and were connected to a common network, due to the uneven sampling rate of Dreem headband there was a small time drift between the two data sets. An example of the uneven sampling rate is depicted in Figure S6. It was noted that the sampling rate of the DHB 3 was more uniform and the drift relative to the PSG was smaller compared to DHB2.

The outline of the synchronisation approach is depicted in the flow diagram Figure S7 [1]. The approach uses first 60 mins of the recorded data from the PSG and the DHB to perform the synchronisation. The sample timestamps available in the hdf file of the DHB recording was used for this purpose. The PSG is interpolated and resampled to the DHB sample timestamps and the timeshift between the DHB and PSG EEG signals are estimated using FFT correlation for all channels. The mode of the timeshift estimates is used to align the DHB data with PSG. The approach provides accurate estimates (N=29, $r^2$=1.00, p<0.001; Bias = -0.88 ± 0.27 s) of time shift compared to the synchronisation package provided by the Dreem Research company for DHB 2 (FigureS8). An example of the first 60 mins of the DHB & PSG recording before and after synchronisation plot is depicted in Figure S9.

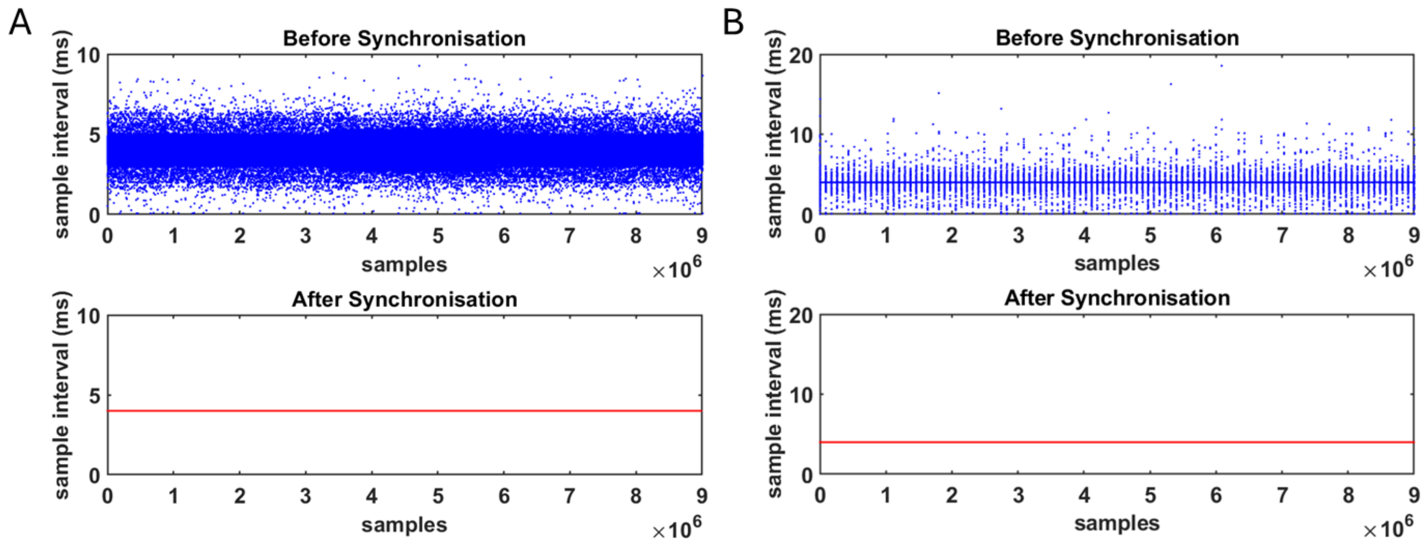


**Figure S6. Examples of uneven sampling intervals in the Dreem headband data and evenly sampled data that is generated through resampling during the synchronisation process. A. Dreem headband 2 and B. Dreem headband 3.**

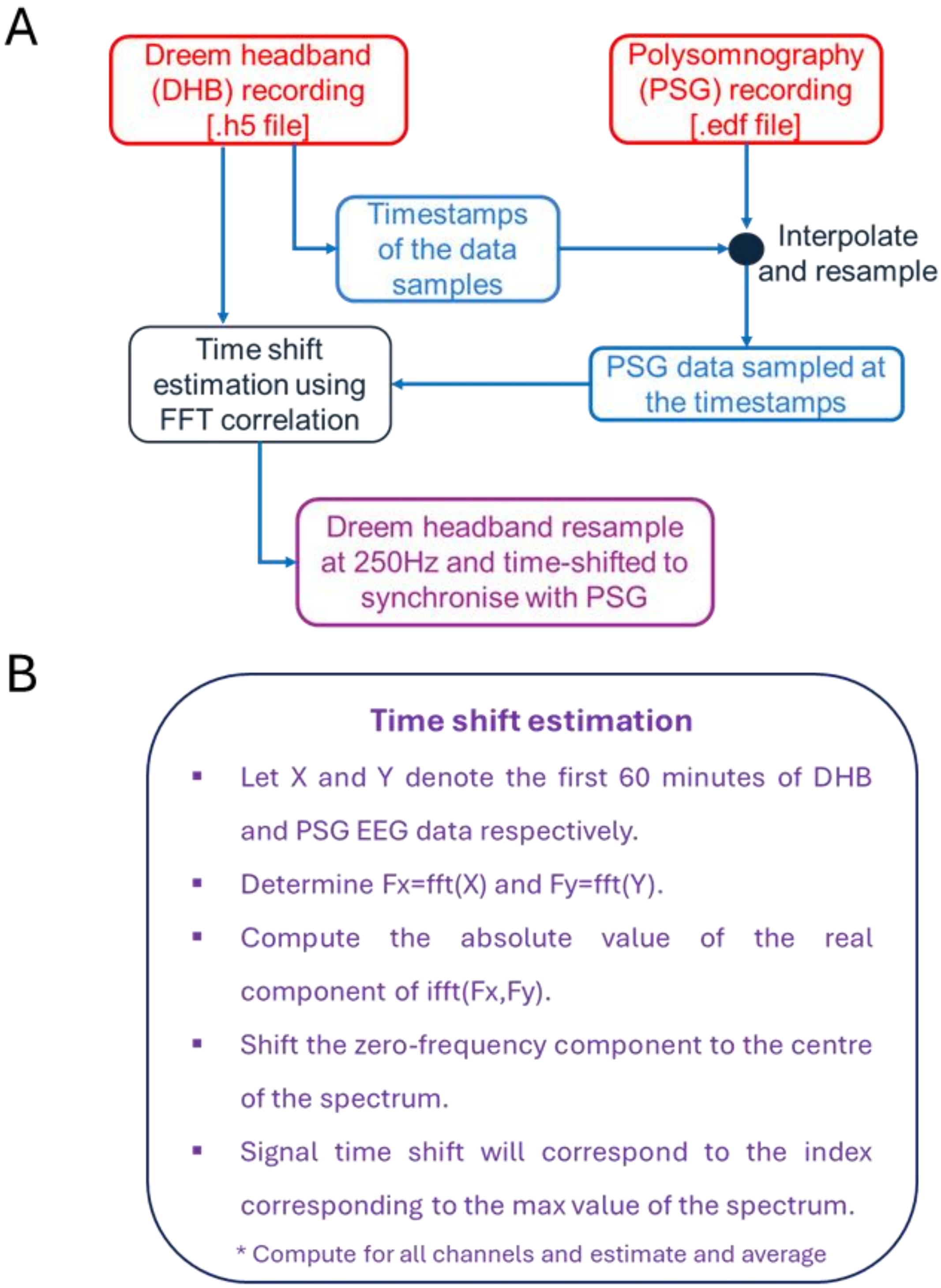


**Figure S7. A. Outline of the synchronisation approach used to align the Dreem headband (DHB) and Polysomnography (PSG) data. B. Method used for estimating the optimal time shift between DHB and PSG to achieve synchronization.**

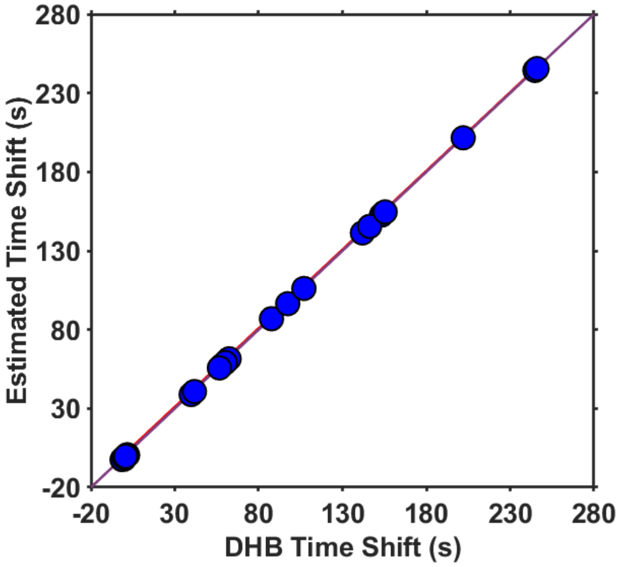

Figure S8. Association between the time shift estimates of the proposed approach and the time shift provided by the synchronisation software provided by Dreem Research (N= 29).

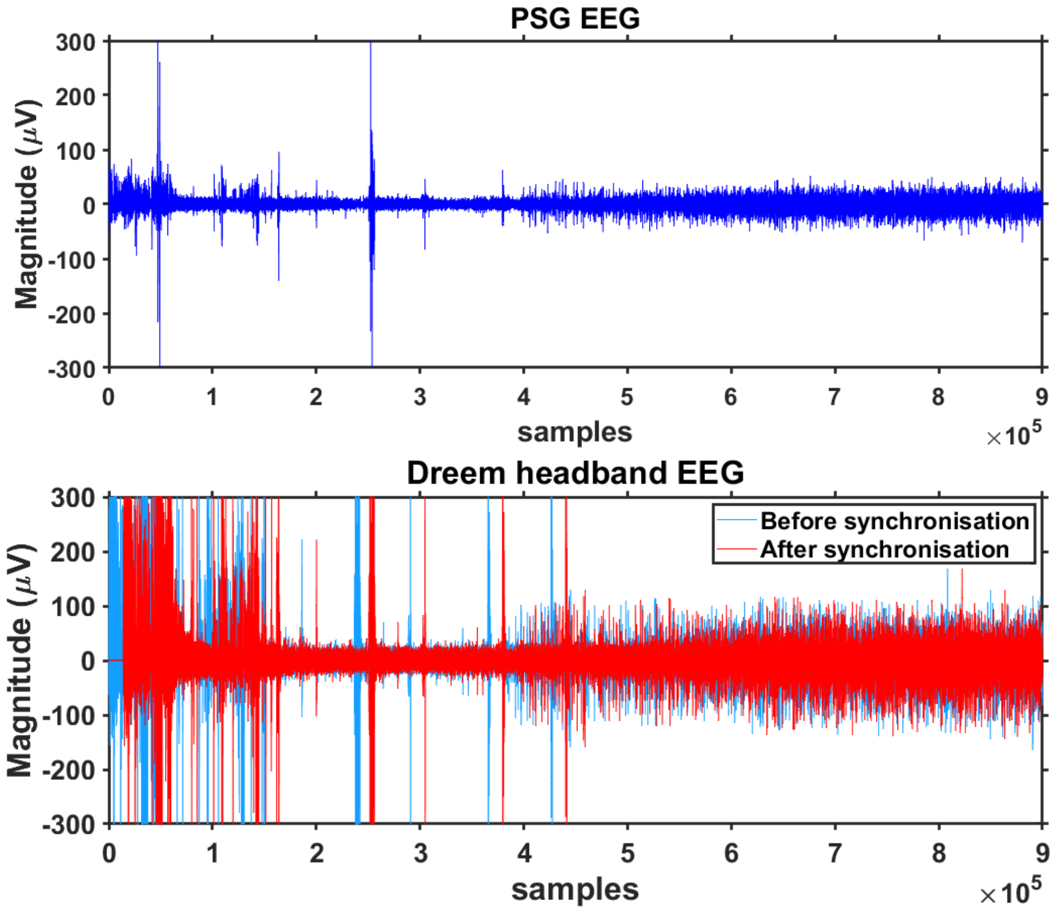

Figure S9. An example of Dreem headband data before and after synchronisation.

### Comparison of the Lights off period

The sleep reports automatically generated by the DHB algorithm contains variables about start and stop times of the recoding and variables named Lights off and Lights on. We compared our experimental PSG lights off and on times with the DHB times. The DHB recording start time had a high correlation with PSG lights off time (N=62, ρ=0.73, p<0.001) while there was no significant correlation between the DHB end times and lights on times (N=62, ρ=0.20, p=0.143). The correlation between the DHB and PSG lights off times were higher (N=62, ρ=0.60, p<0.001) than that of the lights on times (N=62, ρ=0.32, p=0.025) [See Figure S10].

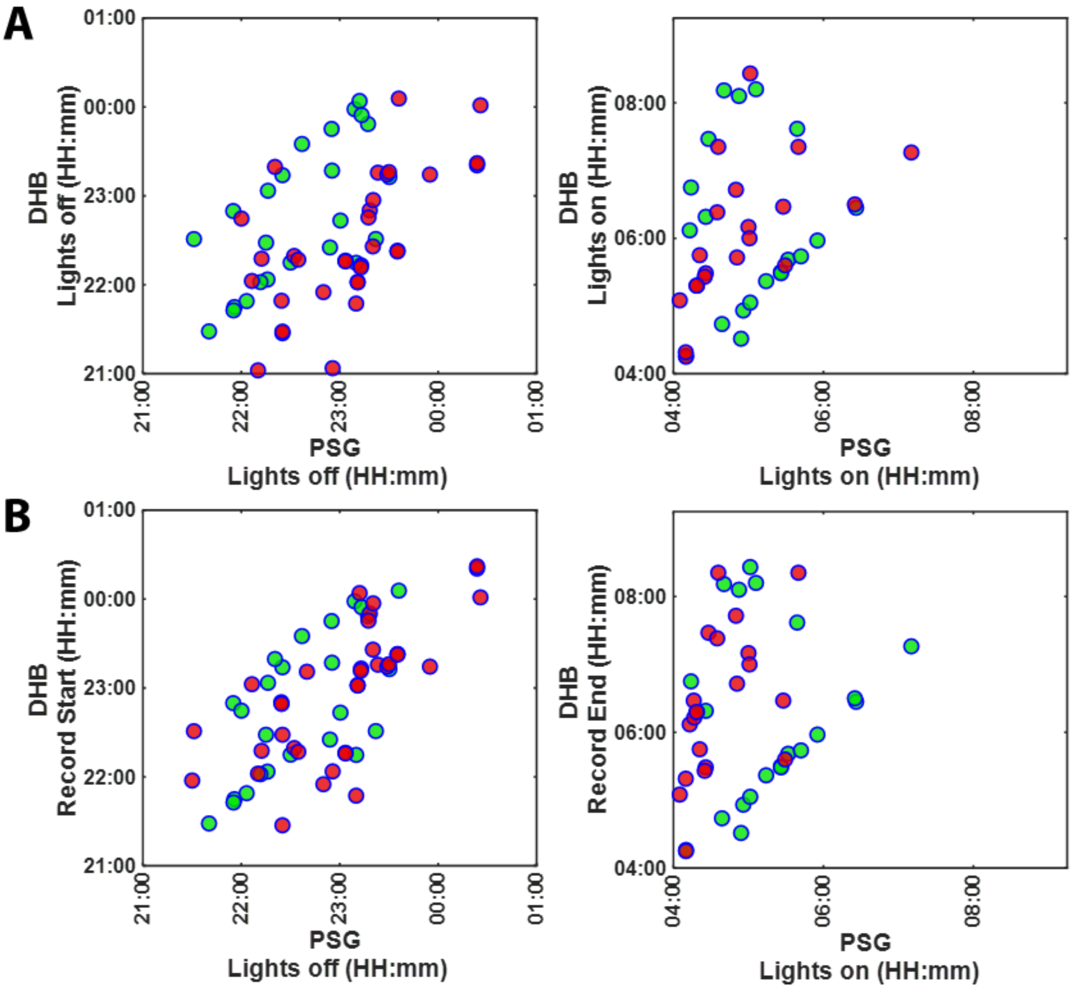


**Figure S10. Comparison of the DHB lights off/on times and recording start/end times with PSG lights off/on times. A. DHB lights off/on and B. DHB Record start/end times. The data points in green indicate DHB 3 estimates.**

#### PSG Lights off period sleep summary definitions

The definitions used for estimating the sleep summary measures from the PSG/device hypnogram in the lights off period (Analysis period manual [AP-M]) is summarised in below.

**Table S5. Sleep summary measure definitions**

| Sleep measure | Definition |
|---|---|
| Total sleep time (TST, min) | time in minutes between lights off to lights on scored as NREM or REM. Wake within the period between lights off and lights on is not included |
| Sleep Onset Latency (SOL, min) | time in minutes from lights off to the first epoch of NREM or REM |
| Wake after sleep onset (WASO, min) | time in minutes of epochs scored as wake from SOL until lights on |
| Total Recording Time (TRT, min) | Time in minutes from lights off to lights on TRT |
| Sleep Efficiency (SEFF, %) | percentage of TST against TRT |
| Sleep stage duration | time in minutes scored as sleep stage (N1, N2, N3, REM, Wake, NREM, Deep sleep, Light sleep) from lights off to lights on |
